# Effectiveness of vaccination against symptomatic and asymptomatic SARS-CoV-2 infection: a systematic review and meta-analysis

**DOI:** 10.1101/2021.08.25.21262529

**Authors:** A. Meggiolaro, M. Sane Schepisi, G. Nikolaidis, D. Mipatrini, A. Siddu, G. Rezza

## Abstract

**OBJECTIVE:** To assess the effectiveness of SARS-CoV-2 vaccines in terms of prevention of disease and transmission. The evaluation was narrowed to two mRNA vaccines and two modified adenovirus vectored vaccines.

**METHODS:** A frequentist random effects meta-analysis was carried out after data extraction. Risk of bias of the included studies was assessed using New-Castle-Ottawa Scale. The overall risk of SARS-CoV-2 infection confirmed by real time Polymerase Chain Reaction (PCR) was estimated in partially and fully vaccinated individuals. The effect size was expressed as Relative Risk (RR) and RRR (RR reduction) of SARS-CoV-2 infection after vaccination. Potential sources of heterogeneity were investigated through between-study heterogeneity analysis and subgroup meta-analysis.

**RESULTS:** The systematic review identified 27 studies eligible for the quantitative synthesis. Partially vaccinated individuals presented a RRR=73% (95%CI=59%-83%) for any positive SARS-CoV-2 PCR (RR=0.27) and a RRR=79% (95%CI=30%-93%) for symptomatic SARS-CoV-2 PCR (RR=0.21). Fully vaccinated individuals showed a RRR=94% (95%CI=88%-98%) for any SARS-CoV-2 positive PCR (RR=0.06) compared to unvaccinated. According to the subgroup meta-analysis, full BNT162b2 vaccination protocol achieved a RRR=84%-94% against any SARS-CoV-2 positive PCR and a RRR=68%-84% against symptomatic positive PCR. The RR for any SARS-CoV-2 positive PCR remained higher within elderly groups aged ≥69 years (RR=0.12-0.15) compared to younger individuals (RR=0.05-0.12). The RR against B.1.351 infection approached 0.40 for any positive PCR and 0.36 for symptomatic SARS-COV-2 while the RR of any B.1.1.7 infection was 0.14.

**CONCLUSION:** The current licensed vaccines may be transmission blocking, especially after full vaccination protocol. Given the substantial heterogeneity, results should be interpreted with caution. Subgroups meta-analyses suggested that the risk of any SARS-CoV-2 infection may be higher for non-B.1.1.7 variants and individuals aged ≥69 years. Further data and longer follow-up are required to investigate additional sources of heterogeneity and the effectiveness of SARS-CoV-2 vaccination within population subgroups.

**STRENGTHS:**

- Real-world data suggest that the current licensed vaccines may be transmission blocking, in particular after full vaccination protocol.
- The risk of any SARS-CoV-2 infection either symptomatic or asymptomatic, may be higher for non-B.1.1.7 variants and individuals aged ≥69 years.

**LIMITATIONS:**

- Given the substantial heterogeneity encountered in this meta-analysis, results should be interpreted with caution
- Fur ther evidence on the impact of SARS-CoV-2 variants are vital in order to monitor mutations associated with vaccine escape

## INTRODUCTION

Since December 2020, infections with SARS-CoV-2 and the respective disease, COVID-19 (Coronavirus Disease 2019), have spread worldwide. On 11 March 2020, WHO declared the COVID-19 outbreak a pandemic. The Coronavirus disease 2019 (COVID-19) has represented a serious threat to public health reporting nearly 4,038,342 deaths and 186,821,815 confirmed cases globally. As of 5 July 2021, 3,114,766,865 vaccine doses have been administered.[1]

Coronaviruses (CoVs), including SARS-CoV, MERS-CoV, and SARSCoV-2, are positive-sense, single stranded RNA viruses with four structural proteins (S protein, envelope (E) protein, membrane (M) protein, and nucleocapsid (N) protein).[2] The spike protein of SARS-CoV-2 binds to ACE2 receptors on target cells and acts as immunodominant antigen, eliciting both antibody and T-cell responses.[3]

Most COVID-19 candidate vaccines have been developed including mRNA, adenoviral vectored vaccines, inactivated virus, and adjuvanted.[4] During the COVID-19 pandemic, the European medicines Agency (EMA) has applied a conditional marketing authorisation for the fast-track approval of safe and effective COVID-19 vaccines in the EU. Initially, EMA has recommended three COVID-19 vaccines to prevent COVID-19: Comirnaty (BNT162b2), Vaxzevria (ChAdOx1/AZD1222), Moderna (mRNA-1273). On March 2021, EMA has further expanded the arsenal of COVID-19 vaccines available to European member states with the COVID-19 vaccine Janssen (Ad26.COV2.S). Each licensed vaccine should meet rigorous standards for safety, efficacy and quality, hence, despite the earlier market authorization, a comprehensive monitoring process is generated post-approval.[5]

The vaccines currently authorised in the EU have proven high efficacy in Phase III clinical trial – such as Moderna’s COVID-19 vaccine with 94.5% efficacy and Pfizer’s with 95% efficacy – and likewise, they are expected to be highly effective in the real world. The effort to measure vaccine effectiveness is crucial to encouraging the vaccine uptake and to improving vaccine technology.[6, 7] While efficacy estimate, in terms of degree to which a vaccine prevents disease and/or transmission, is measured under ideal circumstances such as Controlled Randomized Trials (RCTs), effectiveness refers to an estimate of the vaccine performance in the real world. Effectiveness is generally measured through observational studies where participants are not randomly assigned to an intervention versus a control group.[8]

The COVID-19 vaccination campaign has offered different options and the vaccines’ effect is not necessarily equal.[9-12] The controversy on whether the vaccination is sufficient to prevent any SARS-CoV-2 infection or it merely converts infections from symptomatic to asymptomatic has remained unsolved. Asymptomatic infection following vaccination could still spread the virus and nosocomial COVID-19 infections have been seemingly persistent within health care workers (HCWs), despite the vaccination.[13]

The main objective of this study was to estimate the effectiveness of conditional market licensed COVID-19 vaccines in terms of prevention of disease and transmission. The vaccine effectiveness (VE) was measured as relative risk (RR) and relative risk reduction (RRR) of any SARS-CoV-2 infection, confirmed by polymerase chain reaction (PCR) test, on partially and fully vaccinated individuals respect to unvaccinated individuals. The meta-analysis estimated the overall RR of asymptomatic and symptomatic SARS-CoV-2 infections after partial and full vaccination protocol. The evaluation was narrowed to two mRNA vaccines (Comirnaty and Moderna), a modified adenovirus vaccine (COVID-19 Vaccine AstraZeneca or Vaxzevria) and a recombinant human adenovirus vector (Ad26.COV2.S or Janssen or J&J).

Worldwide mass vaccination requires evidence synthesis on SARS-CoV-2 VE in order to drive health policy strategies. In particular, VE findings may address policy makers’ choices over mitigation measures following mass vaccination campaign, vaccine supply and deployment, vaccination protocol implementation.

## METHODS

### Protocol registration

The systematic review was prospectively registered on PROSPERO (n. CRD42021240143).

### Systematic Review

The systematic review started on March 2021 and was concluded on May 15 2021. The main purpose of this research was to assess the risk of COVID-19 occurrence (any positive RT-PCR test) among vaccinated subjects. Moreover, we aimed at quantifying the risk of developing a symptomatic COVID-19 after vaccination. The evaluation was narrowed to BNT162b2, mRNA-1273, ChAdOx1/AZD1222 and Ad26.COV2.S vaccines. This research followed the PRISMA 2020 statement.[14]

The search strategy was presented in details in Annex 1. We used seven different web engines, including early stage research platforms: Pubmed, Cochrane, clinicaltrial.gov, and COVID-NMA, medRxiv, SSRN, and Authorea. No restriction on language, setting or publication date was imposed.

The papers were selected initially by titles and abstracts. The full texts suitable for the quantitative synthesis were collected in an excel database for the data extraction. The exclusion criteria included lack of suitable data, and study design. We narrowed the quantitative synthesis to non-experimental studies only.

### Data extraction

The data extraction was performed by two authors, independently. The results of the respective phase III RCTs were used as reference for data extraction.[6, 7, 15, 16] The information derived from each full text was classified into four categories of information: 1) outcome, 2) study characteristics (design, publication status, year of publication), 3) participants characteristics (mean age, severity of COVID-19 symptoms, dose of SARS-CoV-2 vaccine, SARS-CoV-2 lineage) and 4) risk of bias. Disagreements were resolved by discussion or, if necessary, consultation with a third party.

#### Outcome

The primary endpoint of this reasearch was to measure the overall relative risk (RR) of any SARS-CoV-2 infection 14 days after the first dose (partially vaccinated), seven days after the second dose (fully vaccinated) and 14 days after at least one dose uptake.

As secondary endpoint, we aimed at measuring the risk of symptomatic infection following SARS-CoV-2 vaccination, and, if possible, quantifying the risk of hospitalization and death after partial and full vaccination protocol.

To this purpose, any COVID-19 infection, either symptomatic or asymptomatic, confirmed through PCR was considered as a COVID-19 case. Studies which did not declare to routinely perform a PCR test after vaccination were excluded. Documented SARS-CoV-2 infections (any positive rtPCR) at the baseline were excluded from the effectiveness analysis.

The risk of infection between vaccinated and unvaccinated groups was calculated as relative risk (RR). In order to quantify how much SARS-CoV-2 vaccination reduced the risk of infection relative to the control group, we computed the relative risk reduction (RRR), according to the formula 100x(1-⍰RR).

#### SARS-CoV-2 infection episodes

The primary analysis included the RR of any new positive infection episode among vaccinated and unvaccinated groups confirmed through PCR test. No restriction was imposed to PCR cycle threshold. The impact of SARS-CoV-2 vaccination on infection severity was investigated in secondary analysis as RR of symptomatic positive PCR, and possibly, RR of hospitalization and death following partial and full vaccination protocol. Both self-reported symptoms and symptoms ascertained through clinical visit were included.

Finally, COVID-19 episodes were classified as compatible with the B.1.1.7 variant and incompatible (i.e. B.135, non-B.1.1.7, unspecified).

#### Experimental group

All adults eligible to undergo SARS-CoV-2 mass vaccination were included in the experimental group. BNT162b2 has been the first vaccine authorized for the emergency rollout, hence, more evidence on effectiveness was expected in the scientific literature. For the definition of case in the vaccinated groups we referred to Polack et al. Phase III RCT.[6] The same rules were applied to ChAdOx1/AZD1222 and mRNA-1273. Concerning ChAdOx1/AZD1222, only few studies on VE after the first dose administration were available, as the vaccine was approved later for the emergency rollout.[15] Ad26.COV2.S single dose administration was considered as full vaccination protocol, although the induction time lasts two weeks (Fig. 1).[16, 47]

**Figure 1).**
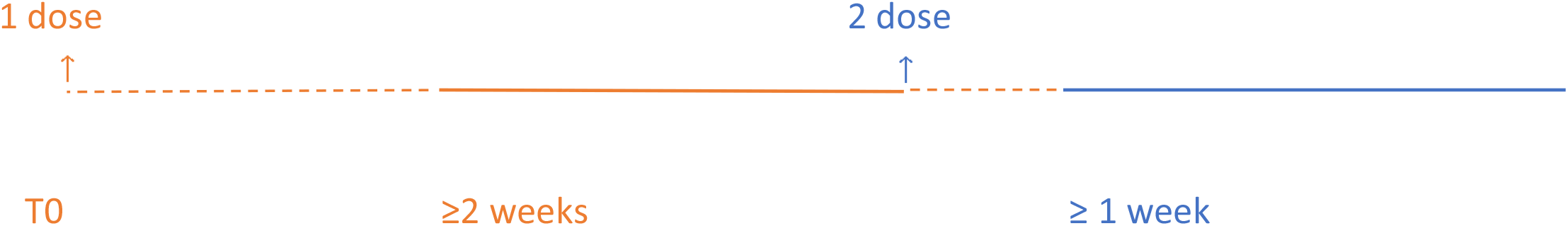
Experimental group follow-up: the dashed line represents the *induction time* for the first and the second vaccine doses respectively. The solid line underpins the follow-up period.

For each vaccine, the 14 days ensuing the first jab were considered as induction time for building COVID-19 immunogenity, therefore, SARS-CoV-2 cases occurred within two weeks from the first dose were not included.[6, 17] In order to assess the effectiveness of full vaccination protocol, the upper follow-up limit was set at seven days after the second dose. (Fig. 1) Only SARS-CoV-2 cases occurred at least one week after the second dose were attributed to the lack of vaccine protection.[6] Moreover, cases occurred within these seven days were still attributed to the first dose effectiveness (Fig. 1).

VE against any positive SARS-COV-2 on at least one dose, was tested after 14 days from the first dose administration.

#### Control group

In the general population, the mean incubation period for COVID-19 symptoms onset was estimated as 5.8 days.[18] Insofar, vaccination has been prioritized towards older and at-risk individuals and this aspect might bias the comparison with the unvaccinated cohort, which generally was younger and healthier. Overall, cohort studies drew the control group from the same population as the vaccinated group, while in few cases the cohort was the same community observed before-after the vaccination. For reasons mostly related to dearth of detailed information on unvaccinated individuals, the beginning of the follow-up for the control group was established at T0 (Fig. 1).

#### Risk of bias

In order to assess the risk of bias, we employed the Newcastle-Ottawa Scale (NOS) in cohort and case-control studies, and the NOS adapted for cross-sectional studies. For systematic review and meta-analysis, we used AMSTAR 2 checklist.[49]

### Statistical analysis

We performed a frequentist meta-analysis using the inverse variance (IV) method.[19] In addition, the Mantel-Haenszel (MH) method was applied because we expected to encounter sparse data, especially in estimating the RR of symptomatic cases after vaccination. The analysis was executed on R (version 4.0.5).

In the early phases of the mass vaccination campaign we expected the selected studies to be statistically heterogeneous. The main assumption underpinning the fixed effect model (FE) is that the true effect size is the same in all studies. This assumption was implausible in this systematic review, as the SARS-CoV-2 vaccination campaign has targeted priority population subgroups in the early stages. Therefore, the random effect (RE) meta-analysis was considered more appropriate to estimate the overall effect size. However, both FE and RE outputs were reported. In order to test for the overall heterogeneity both the Cochran’s Q (chi-squared statistic) and I-squared values (I^2^) were calculated. The Der Simonian and Laird method estimated the between studies variance τ2.[20]

As the between-study heterogeneity can be caused by studies with extreme effect size, low quality or small sample size, the outliers test was implemented in order to detect which studies did not fit in the meta-analysis. Moreover, we explored the robustness of our meta-analyses results using influence analyses.[21] The Baujat Plot displayed which studies overly contributed to the heterogeneity,[22] while the leave-one-out analysis recomputed the pooled effect size and the I^2^ with one study omitted at each step (Annex 3). The Graphic Display of Heterogeneity (GOSH) with Knapp-Hartung adjustment fits the meta-analysis model over all possible subsets of the included studies.[23] Three clustering algorithms detect potential patterns in the data distribution. GOSH analysis allows the identificantion of the studies that contribute to the overall heterogeneity and, possibly, to derive subgroup analysis therefrom (Annex 3).[20]

The subgroups meta-analysis was performed to identify further heterogeneity sources.[24] As we expected a limited number of studies in some subgroups (n<10), we opted for a FE model in subgroup meta-analysis. The subgroup levels were considered fixed and exhaustive. Nevertheless, RE and mixed model were added to the output, in order to catch discrepancies that might yield to inconclusive results (reported in Annex 4.1). In mixed model (or fixed plural model), subgroups are assumed to share a common estimate of the between-study heterogeneity. However, the general approach was to evaluate a separate τ^2^ across subgroups when the number of studies was greater than five, because the estimate of τ^2^ may be imprecise when the number of studies in a subgroup is smaller (i.e. n≤5).[20, 25]

The subgroups meta-analysis enclosed vaccine type, quality, age of vaccinated population, and SARS-CoV-2 lineage detected through PCR tests. Although five or more studies were needed to consistently achieve power from random effects meta-analysis, given a need for a conclusion, two studies were considered sufficient for the statistical inference.[26, 27]

In order to test whether publication biases have influenced the study conclusions we performed a funnel plot (Annex 4.2).[28]

## Results

The web search provided 7,760 unduplicated records. Overall, 31 studies were selected for data extraction and qualitative data synthesis. (Fig. 5, Annex 2). There were 24 cohort studies, two case-control, one cross sectional, one matched observational, and three test negative case-control. Ten studies were performed in UK, eight in US, and five in Israel. RR of SARS-CoV-2 infection among Health care workers (HCWs) was examined by 13 studies; four studies analysed long term care facilities (LCTF) residents and nine the general population. (Tab.3, Annex 2)

### Qualitative synthesis

Four studies met the inclusion criteria for the systematic review but were not included in the quantitative analyses. [50, 53, 54, 55]

A study from Israel assessed the BNT162b2 effectiveness through national surveillance system. The study reported significant VE for asymptomatic SARS-CoV-2 infection (90.4%), symptomatic COVID-19 disease (96.3%), hospitalization (96.0%), severe and critical hospitalization (96.2%), and death (93.3%).[50]

Drury et al. conducted a study through the COVID Symptom Study app in UK. The results showed a progressive risk reduction for SARS-CoV-2 infections among vaccinated over time. Both BNT162b2 (risk reduction increased from 58% to 72%) and ChAdOx1/AZD1222 (risk of infection increased from 39% to 60%) single dose effectiveness was tested against unvaccinated subjects.[53]

In a prospective cohort study conducted among frontline HCWs in US, Thompson et al. reported 90% VE for full immunization and 80% for partial immunization.[54]

In a prospective cohort-study conducted on population registries in Scotland, Vasilieu et al. evaluated the effectiveness of BNT162b2 and ChAdOx1/AZD1222 single dose administration against COVID-19 hospitalization. VE was 85% for BNT162b and 94% for ChAdOx1 /AZD1222.[55]

### Meta-analysis

In the following meta-analysis a RE model with inverse variance (IV) method was used throughout. The evidence synthesis was based on RR as measure of effect. Additionally, the FE estimates as well as the Mantel-Haenszel outputs were reported in Table 1. The between-study variance τ^2^ was computed for each RE meta-analysis.

**Table 1).** Meta-analyses: relative risk of SARS-CoV-2 of positive PCR test after vaccination. Fixed (FE) and Random Effect (RE), inverse variance method (IV) and Mantel-Haenszel (MH) method. RR of any positive PCR and symptomatic positive PCR after SARS-CoV-2 vaccination. Any SARS-CoV-2 vaccination protocol.

| Method | PCR test | SARS-CoV-2 vaccination protocol | N | Fixed effect model | | Random effect model | | $\tau^2$ | I<sup>2</sup> |
| --- | --- | --- | --- | --- | --- | --- | --- | --- | --- |
|  |  |  |  | RR [95%-CI] | p-value | RR [95%-CI] | p-value |  |  |
| Inverse Variance (IV) | any positive PCR | Partially vaccinated | 22 | 0.4115 [0.4025; 0.4207] | 0 | 0.2657 [0.1710; 0.4127] | < 0.0001 | 1.0747 [0.6223; 3.0350] | 99.70% |
|  |  | Fully vaccinated | 17 | 0.1204 [0.1120; 0.1295] | 0 | 0.0586 [0.0266; 0.1292] | < 0.0001 | 1.6228 [1.0654; 2.4896] | 99.00% |
|  |  | At least one dose | 18 | 0.3813 [0.3752; 0.3875] | 0 | 0.1617 [0.1130; 0.2313] | < 0.0001 | 0.5862 [0.4444; 2.6714] | 99.70% |
|  | symptomatic positive PCR test | Partially vaccinated | 9 | 0.4885 [0.4658; 0.5122] | < 0.0001 | 0.2181 [0.0685; 0.6944] | 0.01 | 3.0607 [0.7297; 10.0860] | 99.50% |
|  |  | Fully vaccinated | 8 | 0.2439 [0.2231; 0.2666] | < 0.0001 | 0.0629 [0.0245; 0.1613] | < 0.0001 | 1.7560 [1.1035; 14.6961] | 98.80% |
| Mantel-Haenszel (MH) | any positive PCR | Partially vaccinated | 22 | 0.2490 [0.2436; 0.2546] |  | 0.2656 [0.1617; 0.4363] |  | 1.374 |  |
|  |  | Fully vaccinated | 17 | 0.0447 [0.0419; 0.0477] |  | 0.0586 [0.0228; 0.1505] |  | 3.7986 |  |
|  |  | At least one dose | 18 | 0.3369 [0.3315; 0.3424] |  | 0.1616 [0.1122; 0.2328] |  | 0.6083 |  |
|  | symptomatic positive PCR test | Partially vaccinated | 9 | 0.2758 [0.2644; 0.2878] | 0 | 0.2182 [0.0567; 0.8388] | 0.0267 | 4.1669 | 99.60% |
|  |  | Fully vaccinated | 8 | 0.0811 [0.0752; 0.0874] | 0 | 0.0626 [0.0167; 0.2344] | < 0.0001 | 3.5394 | 99.40% |

Overall, 27 out of 31 studies were included in the meta-analysis. Despite some small sample sizes and few infection cases expected among vaccinated individuals, no study presented zero cells during the data extraction. ChAdOx1/AZD1222 effectiveness was investigated in 4 out of 27 studies,[31, 32, 51, 52] while only one study estimated Ad26.COV2.S effectiveness.[47]

The Newcastle-Ottawa Quality Assessment Scale showed a satisfactory quality, with a median NOS score of six, a minimum of five and a maximum of eight.[29]

#### RR of SARS-CoV-2 infection following vaccination

The meta-analysis on first dose VE against any positive PCR pooled 17 studies and 22 entries. Jones’ analysis spanned over a follow-up of two weeks (A) and, additionally, an extended period of six weeks (B).[30] Bernal et al. evaluated the effectiveness of one and two doses of BNT162b2 vaccine on adults aged ≥ 80 years (A80 and B80) and ≥70 years (B70 and C70) in England. Additionally, the effectiveness of ChAdOx1/AZD1222 single dose (A70) and BNT162b2 one dose at least (C70) were tested on adults aged ≥ 70 years (A70).[31] Shotri et al. estimated the protective effects of BNT162b2 (A) and ChAdOx1/AZD1222 (B) first dose against any SARS-CoV-2 infection.[32] One study from Denmark investigated BNT162b2 VE in two cohorts, namely long-term care facility residents (R) and HCWs (H).[33] Abu Raddad et al. measured the effectiveness of BNT162b2 against B.1.1.7 (A), B.1.351 (B), which became predominant within Qatar in the earliest 2021, and any different (C) SARS-CoV-2 variants.[34] According to the RE meta-analysis results (IV), partially vaccinated showed a RRR=73% of any SARS-CoV-2 positive PCR compared to unvaccinated (RR=0.27). The FE model yielded lower effectiveness estimates after partial vaccination (RR=0.41; RRR=59%). Although both FE and RE (IV) showed a statistically significant result (p< 0.0001), the heterogeneity was considerable (τ^2^=1.08; H = 18.43 [17.52; 19.38]; I^2^ = 99.7%); therefore the true size of the effect remained uncertain (Table 1, Fig. 2a).

**Figure 2).**
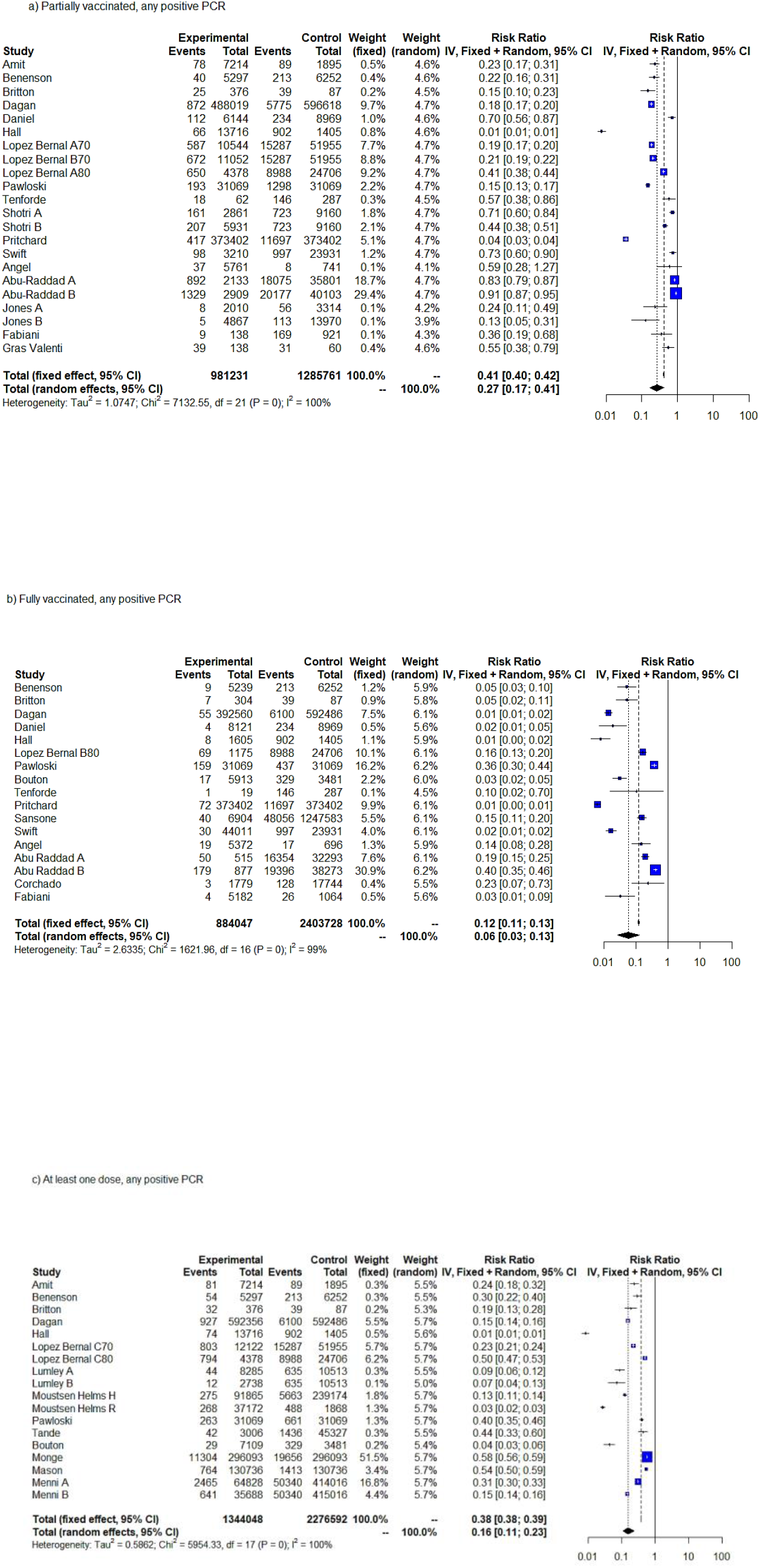
Forest plot, partially vaccinated. Any positive SARS-CoV-2 PCR RR [95%-CI] ≥ 14 days from first dose uptake (a). Forest plot, fully vaccinated. Any positive PCR RR [95%-CI] ≥7 days from full vaccination (b). Forest plot, any positive PCR RR [95%-CI], ≥14 days from vaccination with at least one dose. IV method (c).

In the evaluation of the RR for any positive SARS-CoV-2 PCR following full vaccination protocol, only data on mRNA vaccines (BNT162b2 and mRNA-1273) were available. Sixteen studies and 17 entries were included in this meta-analysis. One study evaluated Ad26.COV2.S effectiveness, whilst “Pritchard” indistinctly assessed the effectiveness of ChAdOx1/AZD1222 and BNT162b2 after full vaccination protocol.[35] In “Abu Raddad” study, data on PCR test were available after two weeks from the BNT162b2 second dose administration.[34] Four studies did not distinguish between BNT162b2 and mRNA-1273 vaccines.[36-39]

Basing on the RE meta-analysis results (IV), the RRR for any positive SARS-CoV-2 PCR following full vaccination protocol was 94% (95%CI=88%-98%) with RR=0.06 compared to unvaccinated individuals (Table 1, Fig. 2b). FE yielded a RRR=88%. Although both FE and RE showed a statistically significant protective effect of full COVID-19 vaccination (p< 0.0001), we should carefully draw conclusion over the true size effect because the heterogeneity was considerable (τ^2^=2.6; H = 10.07 [9.22; 11.00]; I^2^=99.0%). The test for heterogeneity was significant at 1% (Q= 1621.96, p=0). (Table 1, Fig. 2b).

A longer follow-up period represented the main advantage of VE estimation after at least one dose uptake. The mean length of follow-up was indeed 54 days after the first dose administration. The meta-analysis on RR of testing positive for any SARS-CoV-2 PCR included 18 entries and 14 studies. Menni et al. analysed the SARS-CoV-2 infection events on individuals who received one or two doses of BNT162b2 (A) and ChAdOx1/AZD1222 (B).[52] The results of RE meta-analysis (IV) produced a significant RRR=84% for vaccinated individuals compared to unvaccinated, with RR=0.26. The test for heterogeneity was significant at 1% (Q=1030.89, p<0.0001). (Table 1, Fig. 2c)

In order to assess the effectiveness of SARS-CoV-2 vaccination against symptomatic COVID-19 infection, we performed two meta-analyses, in partially and fully vaccinated individuals (Table1). The meta-analysis on full vaccination protocol included only studies estimating VE among individuals aged <69 years. The meta-analyses on VE against symptomatic SARS-CoV-2 infection pooled 9 studies and 17 entries in total. In particular, nine and eight entries pertained to partial and full vaccination protocols respectively. The IV method displayed significant results for both FE and RE (p< 0.05). According to the RE results (IV), partial vaccination achieved a RRR=78% (95%CI= 31%-93%) for symptomatic SARS-CoV-2 infection compared to unvaccinated, while fully vaccinated subjects exhibited a RRR= 94% (95%CI=84%-98%) (Fig. 3a, b). FE estimates of RRR were lower for both partially and fully vaccinated individuals (RRR=51% and RRR=76%, respectively). Both FE and RE were statistically significant (p<0.05).

**Figure 3).**
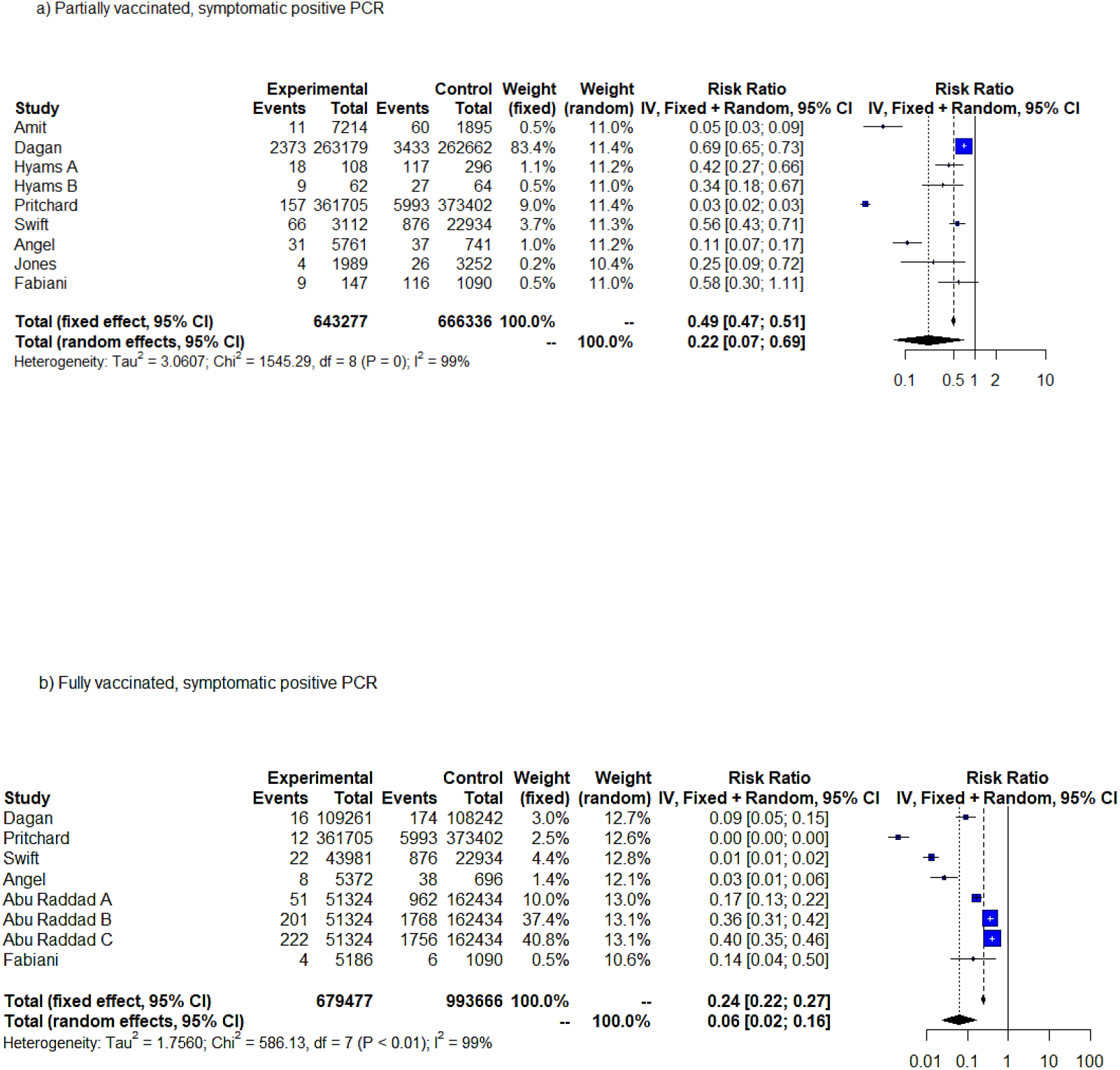
Forest plot, symptomatic positive SARS-CoV-2 PCR RR [95%-CI] ≥ 14 days from first dose uptake (a). Forest plot, fully vaccinated. Symptomatic positive PCR RR [95%-CI] ≥7 days from full vaccination (b). IV method.

Concerning hospitalization risk, a RE meta-analysis over 2 studies,[31, 40] produced a RR=0.38 (95%CI=0.2719; 0.5242) after a full BNT162b2 vaccination protocol. (Q= 2.04, p= 0.1529; I2 = 51.1% [0.0%; 87.6%]). Basing on Bernal et al. data, at least one dose of BNT162b2 vaccine ensured a RRR of death equal to 83%, compared to unvaccinated individuals.[31]

Considering the substantial heterogeneity and the significance of the heterogeneity test, we could not be overly confident that the RR estimate would be robust in every context; therefore, we performed a sensitivity analysis to address the between-study heterogeneity (Annex 3).

#### L’ Abbé plot

The L’ Abbé plots in Fig. 4 confirmed that, overall, infection events were in favour of the control group (unvaccinated). Regarding the meta-analysis on any positive SARS-CoV-2 PCR case after partial vaccination, no study laid above the 45° except “Dagan” and “Angel”, which overlapped the line of no-effect (Fig. 4a), while “Dagan” contributed the most to the overall effect size.[11, 46] Three studies clearly deviated from the effect-size line in Fig. 4a,[34, 38, 41] and this might explain the highest heterogeneity encountered in these data. Four studies were very small and less precise.[38, 41-43] In fully vaccinated meta-analysis (Fig4b), “Pawloski” crossed the no-effect line, while “Abu-Raddad B” and “Hall” evidently deviated from the dashed line of the pooled effect. In Fig. 4c “Hall” and “Lopez Bernal≥80” diverged from the overall RE estimate (light dashed line), while “Monge” appeared as the largest sample sized.[31, 44]

**Fig. 4).**
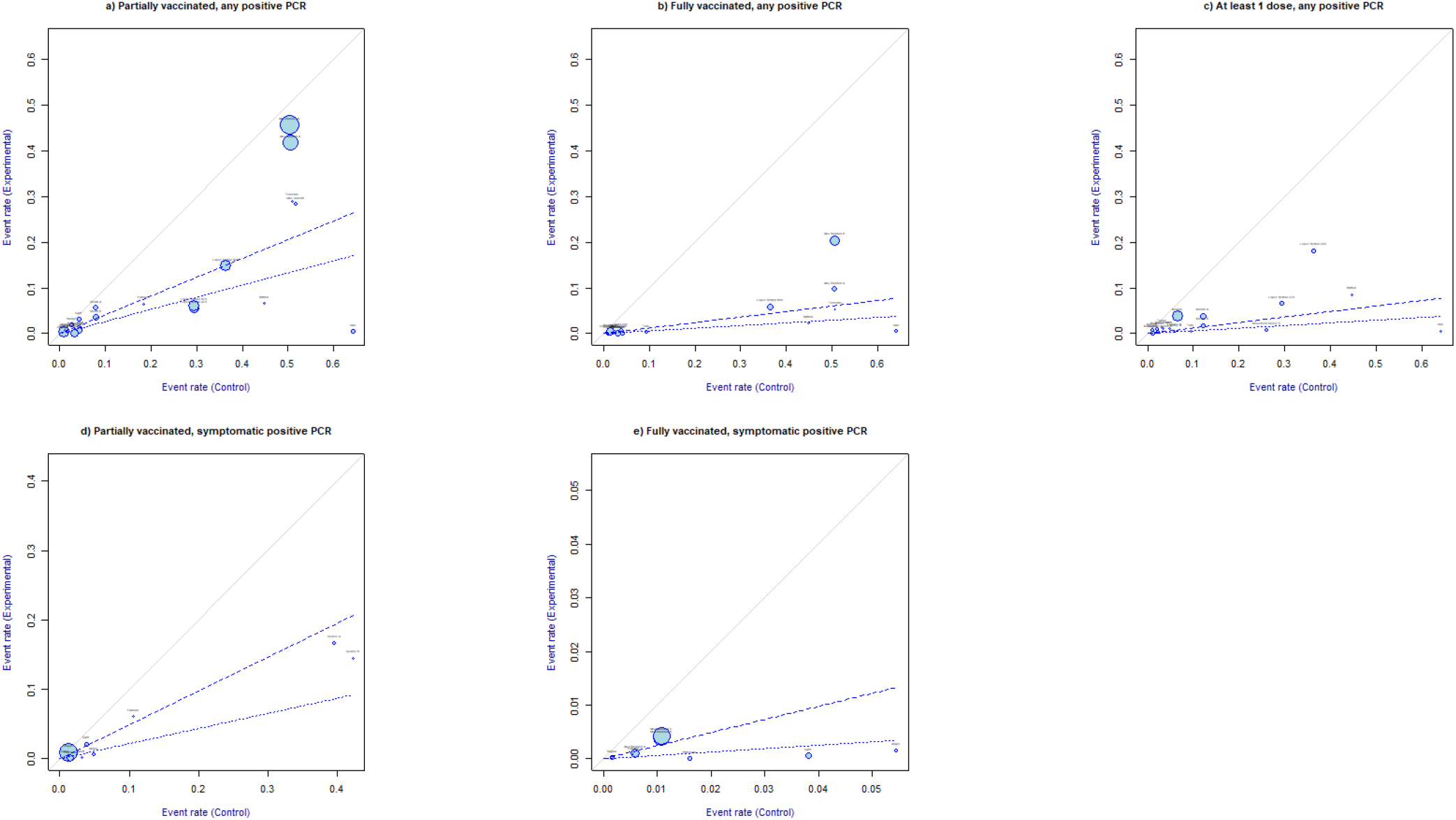
L’Abbè plots. Partially vaccinated, any positive SARS-CoV-2 PCR RR ≥14 days from first SARS-CoV-2 dose administration (a). Fully vaccinated, any positive PCR RR, ≥7 days from full vaccination (b). At least one dose, any positive PCR RR ≥14 days from vaccination with first dose (c). Partially vaccinated, symptomatic positive PCR RR ≥14 days from first dose administration (d). Fully vaccinated, symptomatic positive SARS-CoV-2 PCR RR ≥7 days from full vaccination (e). The sizes of the plotted circles are proportional to the precision of the studies. The dashed lines marked the overall estimate of the log risk for RE (light) and FE (bold). The further the circle from the line of no effect, the greater the difference of event rates between intervention and control arms.

Concerning symptomatic SARS-CoV-2 infections, the L’Abbè plots in Fig. 4d) and 4e) confirmed that the infection rates were greater in unvaccinated groups than partially or fully vaccinated groups. Overall, “Dagan”, “Jones”, and “Abu-Raddad” were the largest sized studies (Fig. 4d and 4e).[11, 13, 45] In Fig. 4d, “Dagan” partially overlapped the 45° line. No study frankly diverged from the light dashed line (RE overall RR) except “Hyams”.[11, 45]

#### Subgroup meta-analysis

The GOSH analysis did not indicate any clear cluster that contributed to the pooled imbalance (Annex 3). In the subgroup meta-analysis, we grouped the studies by type of vaccine, mean age of the sample (≥69 and < 69 years), SARS-CoV-2 lineage, and bias assessment performed through Newcastle-Ottawa Scale (NOS). We classified NOS quality score into satisfactory (NOS ≤6) and good (NOS >6).[29] The meta-analysis was performed over two studies at least.[27] The results of FE model were presented in Tab.2 while mixed and RE subgroup meta-analyses were attached in Tab.10 and 11 (Appendix 4.1). The FE subgroup meta-analysis broadly explained the between-study heterogeneity because all subgroups produced a significant Q test (p< 0.0001). However, considered the residual heterogeneity and the small number of studies in several subgroups, results must be interpreted with caution.

The first subgroup analysis compared different vaccine technologies. Basing on FE subgroup estimates, the RRR after BNT162b2 first dose ranged from 46% to 49% for any positive SARS-COV-2 PCR and from 35% to 32% for symptomatic events. The discrepancy might be explained by the low number of studies included in the subgroup evaluating VE against symptomatic SARS-COV-2 infection (n=6) with respect to the group evaluating the effectiveness of BNT162b2 vaccine against any positive PCR (n=16) (Table 2). Only one study assessed the effectiveness of ChAdOx1/AZD1222 against symptomatic PCR (RR=0.34), however, the effect size was probably overestimated. Corchado et al. analysis suggested a RRR=77% for Ad26.COV2.S,[47] while “Pritchard” did not distinguish between BNT162b2 and ChAdOx1/AZD1222 in VE analysis.[35] In FE the RR estimates were greater within high quality studies (NOS>6), either symptomatic or not (RR=0.12 and RR=0.07 respectively) compared to satisfactory studies (NOS≤6, RR=0.67 and RR=0.48, respectively). Partial vaccination effectiveness appeared lower within studies which examined older population (≥69 years) compared to younger, either on symptomatic PCR (RR= 0.40 and RR= 0.49, respectively) or any positive PCR (RR=0.29 and RR= 0.48, respectively).

**Table 2).** Subgroup meta-analysis, any SARS-CoV-2 protocol. Fixed effect model.

| Subgroups |  | N | Results for subgroups |  |  | Between-groups heterogeneity |  |  |
| --- | --- | --- | --- | --- | --- | --- | --- | --- |
|  |  |  | RR [95%-CI] | I2 | tau2 | Q | df(Q) | p-value |
| a) Partially vaccinated, any positive PCR - fixed effect |  |  |  |  |  |  |  |  |
| Vaccine | BNT162b2 | 16 | 0.5250[0.5123;0.5381] | 99.60% | 0.7549 | 3108.43 | 3 | 0 |
|  | ChAdOx1/AZD 1222 | 2 | 0.2277[0.2122;0.2443] | 98.90% | 0.3565 |  |  |  |
|  | BNT162b2/mRNA-1273 | 3 | 0.2778[0.2474;0.3120] | 98.80% | 1.0692 |  |  |  |
|  | BNT162b2/ChAdOx1/AZD 1222 | 1 | 0.0357[0.0323;0.0393] | -- |  |  |  |  |
| Quality | NOS ≤6 | 15 | 0.4816[0.4704;0.4931] | 99.60% | 0.6113 | 1502.28 | 1 | 0 |
|  | NOS >6 | 7 | 0.1219[0.1142;0.1302] | 99.70% | 3.3748 |  |  |  |
| Age | < 69 years | 15 | 0.4828[0.4702;0.4958] | 99.80% | 1.5669 | 463.24 |  | < 0.0001 |
|  | ≥ 69 years | 7 | 0.2845[0.2733;0.2962] | 98.60% | 0.2424 |  |  |  |
| Lineage | B.1.1.7 | 8 | 0.3903[0.3777;0.4033] | 99.70% | 0.9447 | 3031.61 | 3 | 0 |
|  | B.1.1.7/non B.1.1.7 | 5 | 0.1492[0.1418;0.1569] | 99.70% | 1.4303 |  |  |  |
|  | not specified | 8 | 0.3254[0.2971;0.3565] | 97.10% | 0.6506 |  |  |  |
|  | B.1.351 | 1 | 0.9080[0.8717;0.9458] | -- |  |  |  |  |
| b) Fully vaccinated, any positive PCR - fixed effect |  |  |  |  |  |  |  |  |
| Vaccine | BNT162b2 | 11 | 0.1680 [0.1537; 0.1836] | 98.40% | 1.7775 | 702.84 | 3 | < 0.0001 |
|  | BNT162b2/mRNA-1273 | 4 | 0.1624 [0.1394; 0.1893] | 98.90% | 3.7392 |  |  |  |
|  | BNT162b2/ChAdOx1/AZD1222 | 1 | 0.0062 [0.0049; 0.0078] | -- | -- |  |  |  |
|  | Ad26.COV2.S | 1 | 0.2338 [0.0745; 0.7337] | -- | -- |  |  |  |
| Quality | NOS ≤6 | 11 | 0.1947 [0.1797; 0.2108] | 98.50% | 1.3935 | 827.35 | 1 | < 0.0001 |
|  | NOS >6 | 6 | 0.0112 [0.0094; 0.0134] | 95.90% | 1.6474 |  |  |  |
| Age | < 69 years | 13 | 0.1171 [0.1084; 0.1266] | 99.30% | 3.0726 | 4.21 | 1 | 0.0403 |
|  | ≥ 69 years | 4 | 0.1487 [0.1200; 0.1843] | 65.00% | 0.2931 |  |  |  |
| Lineage | B.1.1.7 | 4 | 0.1448 [0.1231; 0.1703] | 95.90% | 0.817 | 1000.54 | 3 | < 0.0001 |
|  | B.1.1.7/non B.1.1.7 | 5 | 0.0190 [0.0165; 0.0219] | 98.60% | 1.9969 |  |  |  |
|  | not specified | 7 | 0.1692 [0.1452; 0.1971] | 97.70% | 3.4732 |  |  |  |
|  | B.1.351 | 1 | 0.4027 [0.3533; 0.4592] | -- | -- |  |  |  |
| **c) At least one dose, any positive PCR - fixed effect** |  |  |  |  |  |  |  |  |
| Vaccine | BNT162b2 | 12 | 0.2575 [0.2511; 0.2640] | 99.60% | 0.6218 | 2752.22 | 2 | 0 |
|  | BNT162b2/mRNA-1273 | 4 | 0.5641 [0.5517; 0.5767] | 98.50% | 0.4039 |  |  |  |
|  | ChAdOx1/AZD 1222 | 2 | 0.1462 [0.1354; 0.1578] | 83.10% | 0.2114 |  |  |  |
| Quality | NOS ≤6 | 16 | 0.2550 [0.2491; 0.2610] | 99.40% | 0.432 | 2182.95 | 1 | 0 |
|  | NOS >6 | 2 | 0.5525 [0.5402; 0.5650] | 99.90% | 8.9219 |  |  |  |
| Age | < 69 years | 12 | 0.2196 [0.2133; 0.2260] | 99.30% | 0.5076 | 2070.75 | 1 | 0 |
|  | ≥ 69 years | 6 | 0.4927 [0.4832; 0.5025] | 99.80% | 0.5585 |  |  |  |
| Lineage | B.1.1.7 | 4 | 0.3357 [0.3223; 0.3497] | 99.80% | 0.8863 | 1015.58 | 2 | < 0.0001 |
|  | B.1.1.7/non B.1.1.7 | 5 | 0.1463 [0.1372; 0.1560] | 95.00% | 0.3366 |  |  |  |
|  | not specified | 9 | 0.4231 [0.4154; 0.4310] | 99.80% | 0.5975 |  |  |  |
| **d) Partially vaccinated, symptomatic positive PCR - fixed effect** |  |  |  |  |  |  |  |  |
| Vaccine | BNT162b2 | 6 | 0.6572 [0.6245; 0.6915] | 99.60% | 1.1044 | 1414.28 | 3 | < 0.0001 |
|  | ChAdOx1/AZD 1222 | 1 | 0.3441 [0.1763; 0.6715] | -- |  |  |  |  |
|  | BNT162b2/mRNA-1273 | 1 | 0.5552 [0.4336; 0.7111] | -- |  |  |  |  |
|  | BNT162b2/ChAdOx1/AZD 1222 | 1 | 0.0270 [0.0231; 0.0317] | -- |  |  |  |  |
| Quality | NOS ≤6 | 6 | 0.6688 [0.6354; 0.7039] | 99.60% | 0.7447 | 1057.05 | 1 | < 0.0001 |
|  | NOS >6 | 3 | 0.0676 [0.0595; 0.0769] | 99.50% | 3.5694 |  |  |  |
| Age | < 69 years | 7 | 0.4902 [0.4673; 0.5143] | 99.60% | 3.384 | 1.25 | 1 | 0.2636 |
|  | ≥ 69 years | 2 | 0.3962 [0.2736; 0.5737] | 0.00% | -- |  |  |  |
| Lineage | B.1.1.7 | 3 | 0.2410 [0.1801; 0.3224] | 89.10% | 0.5695 | 26.49 | 3 | < 0.0001 |
|  | B.1.1.7/non B.1.1.7 | 2 | 0.5032 [0.4789; 0.5287] | 99.90% | 5.242 |  |  |  |
|  | not specified | 4 | 0.4116 [0.3326; 0.5094] | 94.10% | 1.3278 |  |  |  |
| **e) Fully vaccinated, symptomatic positive PCR - fixed model** |  |  |  |  |  |  |  |  |
| Vaccine | BNT162b2 | 6 | 0.3181 [0.2901; 0.3489] | 94.90% | 0.3245 | 488.27 | 2 | < 0.0001 |
|  | BNT162b2/ChAdOx1/AZD1222 | 1 | 0.0021 [0.0012; 0.0036] | -- | -- |  |  |  |
|  | BNT162b2/mRNA-1273 | 1 | 0.0131 [0.0086; 0.0200] | -- | -- |  |  |  |
| Quality | NOS ≤6 | 5 | 0.3301 [0.3008; 0.3622] | 93.00% | 0.1911 | 492.15 | 1 | < 0.0001 |
|  | NOS >6 | 3 | 0.0085 [0.0063; 0.0116] | 94.60% | 1.4624 |  |  |  |
| Lineage | B.1.1.7 | 2 | 0.1347 [0.1034; 0.1753] | 94.80% | 1.565 | 222.39 | 3 | < 0.0001 |
|  | B.1.1.7/non B.1.1.7 | 3 | 0.2744 [0.2408; 0.3127] | 99.40% | 6.8062 |  |  |  |
|  | not specified | 2 | 0.0166 [0.0111; 0.0248] | 91.80% | 2.5779 |  |  |  |
|  | B.1.351 | 1 | 0.3598 [0.3111; 0.4162] | -- | -- |  |  |  |

Regarding the FE subgroup meta-analysis in fully vaccinated (Table 2), BNT162b2 produced a RRR=83% against any positive PCR and a RRR=68% against symptomatic SARS-CoV-2 infections. The RE and the mixed model produced a RRR=94% against any positive PCR and RRR=84% against symptomatic COVID-19. Further, the FE subgroup meta-analysis relative to the bias assessment (NOS) showed a greater RR for low quality studies with respect to high quality studies (RR=0.01 in symptomatic PCR and RR=0.33 in any positive PCR meta-analyses, respectively). Full vaccination effectiveness against any positive PCR appeared slightly lower within studies which examined older population (≥69 years) compared to younger (RR=0.15 and RR= 0.12, respectively).

Concerning partial vaccination effectiveness against SARS-CoV-2 variants, the RRR of any positive PCR for B.1.1.7 resulted nearly 60% in FE and 81% in mixed model. The RRR increased to 85% in FE and 92% in mixed model after full vaccination. Although only “Abu Raddad” assessed the effectiveness of BNT162b2 against B.1.351 lineage,[34] the RRR was lower for the B.1.351 variant in both partially (RRR=9%) and fully vaccinated subjects (RRR=60%). Regarding symptomatic PCR, the RRR within fully vaccinated individuals remained higher for B.1.1.7 (RRR=86%) with respect to B.1.351 (RRR=64%) (Table 5a, b, e).

#### Publication bias

The funnel plots were displayed in Fig. 26 (Annex 4.2). The contour-enhanced funnel plots did not exhibit publication bias. The majority of the studies fell into p<0.01 except “Tenforde” (Fig. 26 a, d). The Egger test indicated the presence of funnel plot asymmetry in meta-analysis for symptomatic SARS-CoV-2 RR after full vaccination protocol (intercept=-11.271, p=0.026), although it may lack the statistical power to detect bias, because the number of studies was small (n<10).

## DISCUSSION

Despite the considerable between-study heterogeneity, our findings provided evidence that any COVID-19 vaccine is highly effective outside the controlled conditions of the clinical trials. Overall, SARS-CoV-2 vaccination is effective in reducing the number of new COVID-19 events, either asymptomatic or symptomatic, with the greatest benefit achieved after completing the full vaccination protocol. Fewer real-world data were available for ChAdOx1/AZD1222 due to its later regulatory approval. Yet, vaccination with ChAdOx1/AZD1222 resulted in the rare onset of immune thrombotic thrombocytopenia; therefore, it underwent an additional careful monitoring process by European Union (EU) regulatory authorities.[48]

According to our findings, partially vaccinated individuals were only one-fourth as likely (RR=0.26) to develop any SARS-CoV-2 infection, as were unvaccinated individuals; whilst the overall risk of symptomatic infection was slightly lower (RR=0.22). The full vaccination schedule was effective at 94% against asymptomatic and symptomatic positive PCR tests. Concerning hospitalization risk, a meta-analysis over two studies,[31, 40] yielded a RRR=62% ensuing a full BNT162b2 vaccination protocol. Concerning mortality risk, our findings corroborated Bernal et al. study,[31] conducted over 7.5 million adults aged 70 years and older in UK. At least one dose of BNT162b2 was approximately 83%, effective at preventing death compared to unvaccinated individuals, while there was insufficient follow-up to assess ChAdOx1/AZD1222 impact on mortality because of the later rollout.

The subgroup analysis did not find any significant difference in effectiveness between the mRNA and the modified adenovirus vaccines, in part because the evidence, especially for ChAdOx1/AZD1222, was still very scarce. Up to May 15, only two studies evaluated ChAdOx1/AZD1222,[31, 32, 51, 52] and only one Ad26.COV2.S effectiveness.[47] As expected, COVID-19 VE appeared higher in adults aged <69 years. The evaluation of VE against the SARS-CoV-2 variants remained a key point that lacked of robustness in results. In fact, only one study investigated the effectiveness of BNT162b2 against two variants of concern.[34] The RR of infection following a full vaccination protocol appeared larger for B.1.351 (South Africa) variant compared to B.1.1.7 (UK). Although the evidence suggested a lower VE against SARS-COV-2 variants of concern, our findings were not sufficient to assert that COVID-19 full vaccination protocol is 60% effective against B.1.351 and 94% effective against B.1.1.7.

Our research aimed at addressing questions not answered by clinical trials, by including non-randomized studies undertaken after vaccine licensure. Moreover, we attempted to assess VE within different subgroups not included in the preauthorization vaccine clinical trials. Although imprecise, real world data confirmed the experimental evidence. Nevertheless, the lower level of protection assured against emerging variants of concern encouraged to pursue an accurate and robust VE validation.

This systematic review acknowledged several limitations. Undeniably, the expected heterogeneity stemmed from the observational design of the included studies. Unless a randomized process, it is unlikely to be reduced in any circumstances. Moreover, several studies lacked of data on population characteristics. For instance, the meta-analysis predominantly included Caucasian populations whereas vaccines may have different effectiveness across individuals from different ethnicities. Moreover, five studies did not declare to test systematically the vaccinated cohort during the follow-up. The absence of active laboratory surveillance within vaccinated individuals might have resulted in an underestimation of asymptomatic cases. However, we did not consider a different rate of testing as a bias of concern in terms of RR estimate, because both vaccinated and unvaccinated individuals underwent the same case investigation and contact tracing protocol. Moreover, when systematic testing was not performed, asymptomatic testing was available to different extent: workplace exposures (HCWs), out-of-state travellers or per-request.[37] In general, SARS-CoV-2 PCR testing was highly accessible and free of charge.[11, 50]

However, asymptomatic cases which progressed into symptomatic disease could act as source of bias in both vaccinated and unvaccinated groups.[37, 43] Finally, public health mitigation measures might contribute to the underestimation of SARS-CoV-2 vaccine effectiveness. Unfortunately, the extent to which primary prevention restrictions affected VE was beyond the scope of this study.

By way of conclusion, we can state that significant reduction in RR of asymptomatic infection within partially vaccinated was not corroborated by sufficient statistical robustness for the generalizability of results. However, full vaccination effectiveness over symptomatic and asymptomatic SARS-CoV-2 infections risk, confirmed the RCTs results, except larger CI.

To the best of our knowledge, this is the first evidence-based data synthesis on SARS-CoV-2 VE. Our findings support the maximization of the full vaccination coverage and encourage a better calibration of the statistical inference as the mass vaccination wears on. Long-term vaccine effectiveness remains unexplained and needs further investigation. Better evidence over the impact of SARS-CoV-2 variants are vital in order to monitor mutations associated with vaccine escape.

## Supporting information

Annex

## Data Availability

This manuscript does not contain personal and/or medical information about identifiable individuals. The study protocol is available at PROSPERO web registry (n. CRD42021240143). The statistical analysis plan is available upon request from the Authors.

## ETHICS APPROVAL STATEMENT

No ethics committee approval was required.

## CONTRIBUTORSHIP STATEMENT

Each named author has substantially contributed to conducting the underlying research and writing the paper.

## DATA SHARING STATEMENT

this manuscript does not contain personal and/or medical information about identifiable individuals. The study protocol is available at PROSPERO web registry (n. CRD42021240143). The statistical analysis plan is available upon request from the Authors.

## PATIENT AND PUBLIC INVOLEMENT

patients were involved by primary studies.

## Web appendix

Supplementary material

