## Supplementary material for "Effectiveness of vaccination against symptomatic and asymptomatic SARS-CoV-2 infection: a systematic review and meta-analysis": Annex

### Annex 1

### Full Search Strategy

In order to frame the main research question, we applied the PICO (Participant-Intervention-Comparator-Outcomes) framework to define our research question. The search terms were arranged using the boolean logic.

P=population

I=vaccine

C=controls (unvaccinated)

O=effectiveness in CoVID19 prevention (RR)

Search terms: COVID19 vaccine [AND] effectiveness [OR] efficacy

**Last conducted on: 11/05/2021**

**1. PubMed**

(((effectiveness OR efficacy OR Real World OR Phase 3 OR Phase 4)) AND (("Vaccination"[Mesh] OR "Immunization Programs"[Mesh] OR "Vaccines"[Mesh] OR vaccin*[tiab] OR immunization*[tiab] OR immunisation*[tiab]))) AND ((("COVID-19"[Mesh] OR "SARS-CoV-2"[Mesh] OR "COVID-19 Vaccines"[Mesh] OR "COVID-19 Serological Testing"[Mesh] OR "COVID-19 Nucleic Acid Testing"[Mesh] OR "SARS-CoV-2 variants" [Supplementary Concept] OR "COVID-19 drug treatment" [Supplementary Concept] OR "COVID-19 serotherapy" [Supplementary Concept] OR "2019-nCoV" OR "2019nCoV" OR "cov 2" OR "Covid-19" OR "sars coronavirus 2" OR "sars cov 2" OR "SARS-CoV-2" OR "severe acute respiratory syndrome coronavirus 2" OR "coronavirus 2" OR "COVID 19" OR "COVID-19" OR "2019 ncov" OR "2019nCoV" OR "corona virus disease 2019" OR "cov2" OR "COVID-19" OR "COVID19" OR "nCov 2019" OR "nCoV" OR "new corona virus" OR "new coronaviruses" OR "novel corona virus" OR "novel coronaviruses" OR "SARS Coronavirus 2" OR "SARS2" OR "SARS-COV-2" OR "Severe Acute Respiratory Syndrome Coronavirus 2")))

| Search | Query | Results |
|---|---|---|
| #1 | Search: (("COVID-19"[Mesh] OR "SARS-CoV-2"[Mesh] OR "COVID-19 Vaccines"[Mesh] OR "COVID-19 Serological Testing"[Mesh] OR "COVID-19 Nucleic Acid Testing"[Mesh] OR "SARS-CoV-2 variants" [Supplementary Concept] OR "COVID-19 drug treatment" [Supplementary Concept] OR "COVID-19 serotherapy" [Supplementary Concept] OR "2019-nCoV" OR "2019nCoV" OR "cov 2" OR "Covid-19" OR "sars coronavirus 2" OR "sars cov 2" OR "SARS-CoV-2" OR "severe acute respiratory syndrome coronavirus 2" OR "coronavirus 2" OR "COVID 19" OR "COVID-19" OR "2019 ncov" OR "2019nCoV" OR "corona virus disease 2019" OR "cov2" OR "COVID-19" OR "COVID19" OR "nCov 2019" OR "nCoV" OR "new corona virus" OR "new coronaviruses" OR "novel corona virus" OR "novel coronaviruses" OR "SARS Coronavirus 2" OR "SARS2" OR "SARS-COV-2" OR "Severe Acute Respiratory Syndrome Coronavirus 2")) | 130,245 |
| #2 | Search: ("Vaccination"[Mesh] OR "Immunization Programs"[Mesh] OR "Vaccines"[Mesh] OR vaccin*[tiab] OR immunization*[tiab] OR immunisation*[tiab]) | 447,082 |
| #3 | Search: (effectiveness OR efficacy OR Real World OR Phase 3 OR Phase 4) | 10,214,491 |

| | | |
|---|---|---|
| #4 | Search: #1 AND #2 AND #3 Filters: Clinical Trial, Meta-Analysis, Randomized Controlled Trial, Systematic Review, Review | 4,903 |

**2. MedRxiv**

https://www.medrxiv.org/search/%2528effectiveness%252Bor%252Befficacy%252Bor%252Breal%252Bworld%252Bor%252Bphase%252B3%252Bor%252Bphase%252B4%2529%252Band%252B%2528covid%252Bor%252Bsars-cov-2%2529%252Band%252Bvaccine

| Search | Query | Results |
|---|---|---|
| | "(covid-19 or sars-cov-2) and vaccine and (effectiveness OR efficacy OR Real World OR Phase 3 OR Phase 4)" | 2,390 |

**3. SSRN**

https://papers.ssrn.com/sol3/results.cfm?RequestTimeout=50000000

| Search | Query | Results |
|---|---|---|
| | covid-19 and vaccine and effectiveness | 40 |

**4. Authorea**

https://www.authorea.com/preprints

| Search | Query | Results |
|---|---|---|
| | VACCINE + SARS-COV-2 | 7 |

**5. Clinical Trials**

https://clinicaltrials.gov/ct2/results?term=vaccine+%28effectiveness+or+efficacy%29&cond=covid+19+or+sars-cov-2&Search=Clear&age_v=&gndr=&type=&rslt=

| Search | Query | Results |
|---|---|---|
| | (covid-19 or sars-cov-2) AND vaccine AND (effectiveness or efficacy) | 186 |

**6. Cochrane Library**

https://www.cochranelibrary.com/advanced-search?cookiesEnabled

| Search | Query | Results |
|---|---|---|
| | (covid19 vaccine) or (sars-cov-2 vaccine) and effectiveness | 201 |

**7. COVID-NMA**

https://covid-nma.com/vaccines/mapping/

| Search | Query | Results |
|---|---|---|
| | vaccines | 33 |

#### Annex 2

**Fig.5) PRISMA** Flow Diagram

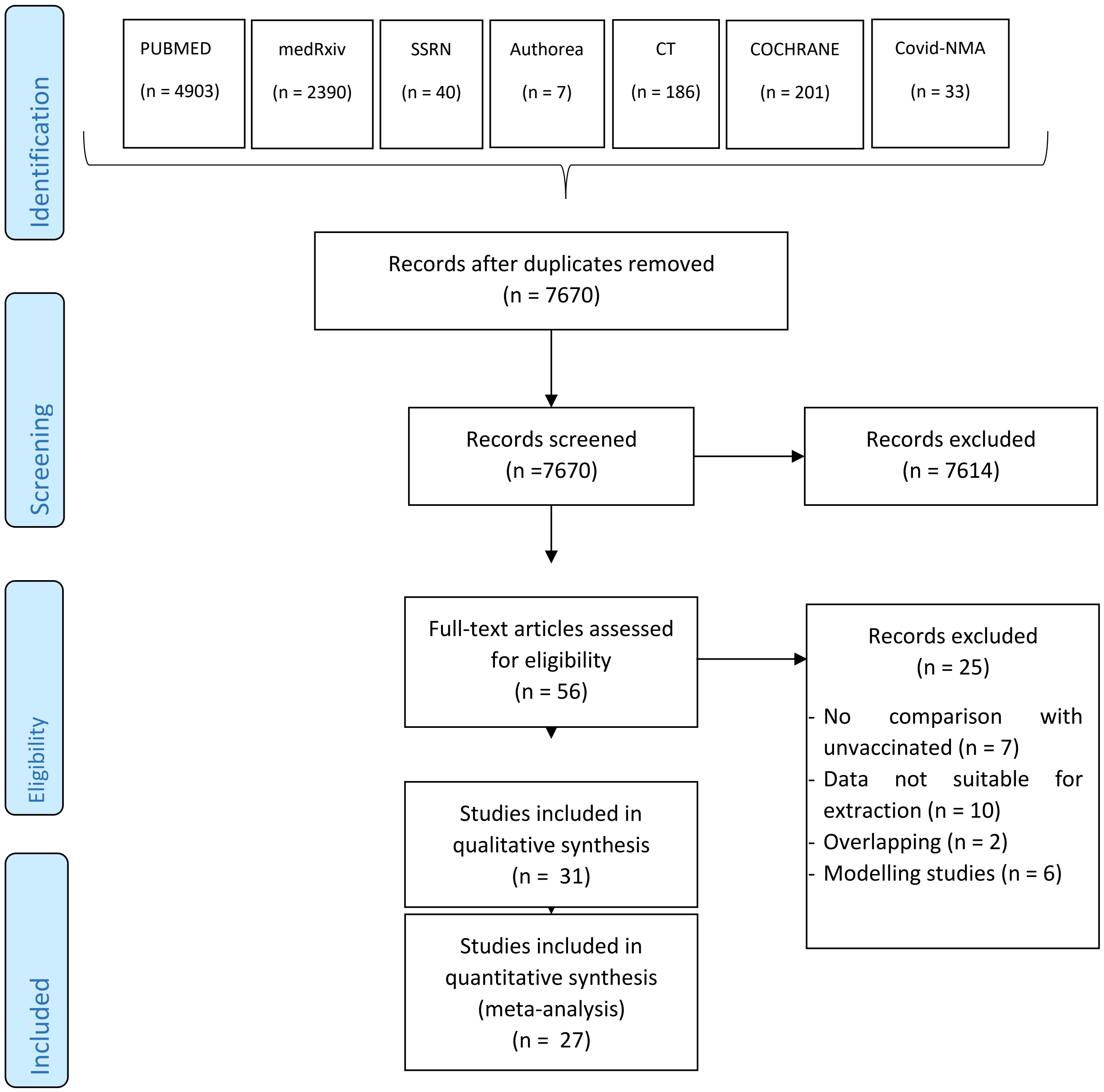

**Table 3)** Characteristics of 31 selected studies**.**

| N | First Author | Country | Study Design | Vaccine | Population (N) | Population type |
|---|---|---|---|---|---|---|
| 1 | **Abu-Raddad** | Qatar | Test Negative-Case–Control | Comirnaty (BNT162b2) | 3,734 | General population |
| 2 | **Amit** | Israel | Retrospective Cohort | Comirnaty (BNT162b2) | 9,109 | Healthcare workers |
| 3 | **Angel** | Israel | Retrospective Cohort | Comirnaty (BNT162b2) | 6,710 | Healthcare workers |
| 4 | **Benenson** | Israel | Retrospective Cohort | Comirnaty (BNT162b2) | 6,252 | Healthcare workers |
| 5 | **Britton** | USA | Retrospective Cohort | Comirnaty (BNT162b2) | 463 | Residents of long-term care facilities |
| 6 | **Corchado** | USA | Matched Cohort | Janssen (Ad26.COV2.S) | 24,145 | General population |
| 7 | **Dagan** | Israel | Matched Observational | Comirnaty (BNT162b2) | 1,193,236 | General population |
| 8 | **Daniel** | USA | Retrospective Cohort | Comirnaty (BNT162b2) Moderna (mRNA-1273) | 23,234 | General population (symptomatic) |
| 9 | **Druri** | UK | Cohort | Comirnaty (BNT162b2), Vaxzevria (ChAdOx1/AZD1222) | 627,383 | General population |
| 10 | **Fabiani** | Italy | Retrospective Cohort | Comirnaty (BNT162b2) | 6,423 | Healthcare workers |
| 11 | **Gras Valenti** | Spain | Case-Control Study | Comirnaty (BNT162b2) | 268 | Healthcare workers |
| 12 | **Haas** | Israel | Cohort | Comirnaty (BNT162b2) | 281,903 | General population |
| 13 | **Hall** | UK | Cohort | Comirnaty (BNT162b2) | 23,324 | Healthcare workers |
| 14 | **Hyams** | UK | Test Negative Case Control | Comirnaty (BNT162b2), Vaxzevria (ChAdOx1/AZD1222) | 434 | Elderly patients |
| 15 | **Jones** | UK | Cross-Sectional | Comirnaty (BNT162b2) | 8,819 | Healthcare workers |
| 16 | **Lopez-Bernal** | UK | Test Negative Case Control | Comirnaty (BNT162b2), Vaxzevria (ChAdOx1/AZD1222) | 156,930 | Elderly population |
| 17 | **Lumley** | UK | Cohort | Comirnaty (BNT162b2) | 23,411 | Healthcare workers |
| 18 | **Mason** | UK | Cohort | Comirnaty (BNT162b2) | 301,461 | Elderly population |
| 19 | **Menni** | UK | Prospective Observational | Comirnaty (BNT162b2), Vaxzevria (ChAdOx1/AZD1222) | 627,383 | General population |
| 20 | **Monge** | Spain | Cohort | Comirnaty (BNT162b2) | 299,209 | Residents in long-term care facilities (LTCF) |
| 21 | **Mouststen-Helms** | Denmark | Retrospective Cohort | Comirnaty (BNT162b2) | 370,079 | Residents in long-term care facilities (LTCF) and Healthcare workers |
| 22 | **Pawlosky** | USA | Retrospective Cohort | Comirnaty (BNT162b2), Moderna (mRNA-1273) | 62,138 | Residents in long-term care facilities (LTCF) and Healthcare workers |
| 23 | **Pritchard** | UK | Cohort | Comirnaty (BNT162b2), Vaxzevria (ChAdOx1/AZD1222) | 373,402 | General population |
| 24 | **Sansone** | Italy | Cohort | Comirnaty (BNT162b2) | 6,904 | Healthcare workers |
| 25 | **Shotri** | UK | Cohort | Comirnaty (BNT162b2), Vaxzevria (ChAdOx1/AZD1222) | 10,412 | Residents of Long-Term Care Facilities (LTCF) |
| 26 | **Swift** | USA | Cohort | Comirnaty (BNT162b2), Moderna (mRNA-1273) | 71,152 | Healthcare workers |
| 27 | **Tande** | USA | Retrospective Cohort | Comirnaty (BNT162b2) | 39,156 | General propulation |
| 28 | **Tara** | USA | Cohort | Comirnaty (BNT162b2) Moderna | 7,109 | Healthcare workers |
| 29 | **Tenforde** | USA | Case-Control | Comirnaty (BNT162b2), Moderna (mRNA-1273) | 417 | Elderly patients |
| 30 | **Thompson** | USA | Cohort | Comirnaty (BNT162b2) Moderna (mRNA-1273) | 3,950 | Healthcare workers |
| 31 | **Vasileious** | Scotland | Cohort | Comirnaty (BNT162b2), Vaxzevria (ChAdOx1/AZD1222) | 4,409,611 | General population |

**Table 4)** Quality assessment Newcastle Ottawa Scale (NOS).

| n | Author | Selection | Comparability | Outcome | Exposure | Total score |
|---|---|---|---|---|---|---|
| 1 | Abu-Raddad | 3 | 1 | 2 | | 6 |
| 2 | Amit | 2 | 1 | 2 | | 5 |
| 3 | Angel | 4 | 2 | 2 | | 8 |
| 4 | Benenson | 2 | 1 | 2 | | 5 |
| 5 | Britton | 2 | 1 | 2 | | 5 |
| 6 | Corchado | 4 | 2 | 1 | | 7 |
| 7 | Dagan | 3 | 1 | 2 | | 6 |
| 8 | Daniel | 2 | 1 | 2 | | 5 |
| 9 | Druri | 3 | 1 | 2 | | 6 |
| 10 | Fabiani | 3 | 1 | 2 | | 6 |
| 11 | Gras Valenti | 2 | 2 | | 2 | 6 |
| 12 | Haas | 3 | 1 | 2 | | 6 |
| 13 | Hall | 4 | 1 | 2 | | 7 |
| 14 | Hyams | 3 | 1 | | 2 | 6 |
| 15 | Jones | 2 | 1 | 2 | | 5 |
| 16 | Lopez-Bernal | 1 | 1 | | 2 | 6 |
| 17 | Lumley | 3 | 1 | 2 | | 6 |
| 18 | Mason | 3 | 1 | 2 | | 6 |
| 19 | Menni | 3 | 1 | 2 | | 6 |
| 20 | Monge | 3 | 1 | 3 | | 7 |
| 21 | Mouststen-Helms | 3 | 1 | 2 | | 6 |
| 22 | Pawlosky | 3 | 1 | 2 | | 6 |
| 23 | Pritchard | 4 | 2 | 2 | | 8 |
| 24 | Sansone | 2 | 0 | 1 | | 3 |
| 25 | Shotri | 4 | 1 | 2 | | 7 |
| 26 | Swift | 3 | 1 | 3 | | 7 |
| 27 | Tande | 3 | 1 | 2 | | 6 |
| 28 | Tara | 3 | 1 | 1 | | 5 |
| 29 | Tenforde | 2 | 2 | | 2 | 6 |
| 30 | Thompson | 2 | 1 | 2 | | 5 |
| 31 | Vasileious | 3 | 1 | 2 | | 6 |

### Annex 3
### Meta-analysis: between-study heterogeneity

#### *A3.1 Partially vaccinated, first dose effectiveness, any Sars-Cov2 positive PCR*

**Table 5) Outliers, partially vaccinated.** The outliers test identified which study's confidence interval did not overlap with the confidence interval of the random pooled effect. Concerning the meta-analysis on partially vaccinated (any positive PCR), seven studies presented an extreme effect size estimate.

| **Identified outliers (random-effects model)** | | | | | | |
|---|---|---|---|---|---|---|
| "Daniel", "Hall", "Shotri A", "Pritchard", "Swift", "Abu-Raddad", "Abu-Raddad" | | | | | | |
| **Results with outliers removed** | | | | | | |
| | **RR [95%-CI %]** | | | **W(fixed) %** | **W(random)** | **exclude** |
| **Amit** | 0.2302 | [0.1706; | 0.3107] | 1.3 | 7.2 | |
| **Benenson** | 0.2217 | [0.1584; | 0.3101] | 1.0 | 7.0 | |
| **Britton** | 0.1483 | [0.0951; | 0.2314] | 0.6 | 6.3 | |
| **Dagan** | 0.1846 | [0.1719; | 0.1982] | 23.1 | 8.2 | |
| **Daniel** | 0.6987 | [0.5591; | 0.8731] | 0.0 | 0.0 | * |
| **Hall** | 0.0075 | [0.0059; | 0.0096] | 0.0 | 0.0 | * |
| **Lopez Bernal A70** | 0.1892 | [0.1747; | 0.2049] | 18.3 | 8.2 | |
| **Lopez Bernal B70** | 0.2066 | [0.1918; | 0.2226] | 21.0 | 8.2 | |
| **Lopez Bernal A80** | 0.4081 | [0.3794; | 0.4389] | 22.0 | 8.2 | |
| **Pawloski** | 0.1487 | [0.1279; | 0.1728] | 5.2 | 8.0 | |
| **Tenforde** | 0.5707 | [0.3805; | 0.8560] | 0.7 | 6.6 | |
| **Shotri A** | 0.7130 | [0.6042; | 0.8413] | 0.0 | 0.0 | * |
| **Shotri B** | 0.4422 | [0.3802; | 0.5143] | 5.1 | 8.0 | |
| **Pritchard** | 0.0357 | [0.0323; | 0.0393] | 0.0 | 0.0 | * |
| **Swift** | 0.7328 | [0.5975; | 0.8988] | 0.0 | 0.0 | * |
| **Angel** | 0.5949 | [0.2781; | 1.2725] | 0.2 | 4.3 | |
| **Abu Raddad** | 0.8283 | [0.7870; | 0.8717] | 0.0 | 0.0 | * |
| **Abu Raddad** | 0.9080 | [0.8717; | 0.9458] | 0.0 | 0.0 | * |
| **Jones A** | 0.2355 | [0.1125; | 0.4930] | 0.2 | 4.4 | |
| **Jones B** | 0.1270 | [0.0519; | 0.3109] | 0.1 | 3.7 | |
| **Fabiani** | 0.3554 | [0.1863; | 0.6782] | 0.3 | 5.0 | |
| **Gras Valenti** | 0.5470 | [0.3811; | 0.7850] | 0.9 | 6.8 | |
| **Meta-analysis with outliers removed** | | | | | | |
| Number of studies combined: k = 15 | | | | | | |
| | **RR [95%-CI %]** | | | **z** | **p-value** | |
| Fixed effect model | 0.2397 | [0.2317; | 0.2481] | -81.99 | 0 | |
| Random effects model | 0.2679 | [0.2130; | 0.3369] | -11.27 | < 0.0001 | |
| **Quantifying heterogeneity:** | | | | | | |
| tau^2 = 0.1657 [0.0822; 0.6127]; tau = 0.4070 [0.2867; 0.7827] | | | | | | |
| I^2 = 97.0% [96.0%; 97.7%]; H = 5.73 [5.00; 6.56] | | | | | | |
| Test of heterogeneity: | | **Q** | **d.f.** | **p-value** | | |
| | | 459.09 | 14 | < 0.0001 | | |
| **Details on meta-analytical method:** | | | | | | |
| - Inverse variance method | | | | | | |
| - DerSimonian-Laird estimator for tau^2 | | | | | | |
| - Jackson method for confidence interval of tau^2 and tau | | | | | | |

**Fig.6)** Forest plot, partially vaccinated. Any positive PCR RR ≥14 days after first dose. Outliers excluded, RE IV method. Identified outliers: "Daniel", "Hall", "Shotri A", "Pritchard", "Swift", "Abu-Raddad A", "Abu-Raddad B". The outlying studies were still displayed. However, their weight in the meta-analysis has been set to 0%; therefore, they were excluded from pooling. Overall RR=0.27 and RRR=73%. Heterogeneity: $\tau^2$= 0.1657 [0.0822; 0.6127]; tau = 0.4070 [0.2867; 0.7827]; $I^2$ = 97.0% [96.0%; 97.7%]; H = 5.73 [5.00; 6.56]. Test of heterogeneity: Q=459.09 (d.f.= 14 p-value < 0.0001). The updated RE RR was 0.23[0.2130; 0.3369], with RRR=77% compared to unvaccinated.

Partially vaccinated, any positive PCR, outliers excluded

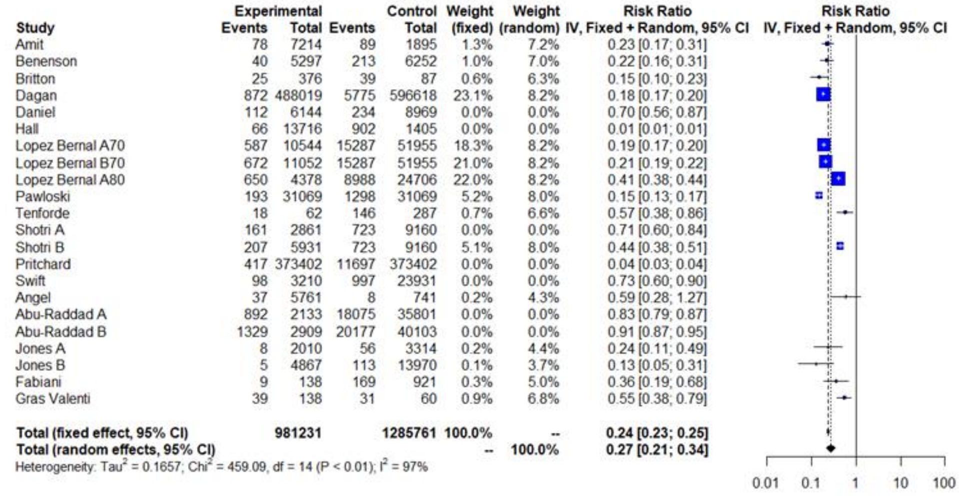


| Study | Experimental Events | Experimental Total | Control Events | Control Total | Weight (fixed) | Weight (random) | Risk Ratio IV, Fixed + Random, 95% CI |
|---|---|---|---|---|---|---|---|
| Amit | 78 | 7214 | 89 | 1895 | 1.3% | 7.2% | 0.23 [0.17; 0.31] |
| Benenson | 40 | 5297 | 213 | 6252 | 1.0% | 7.0% | 0.22 [0.16; 0.31] |
| Britton | 25 | 376 | 39 | 87 | 0.6% | 6.3% | 0.15 [0.10; 0.23] |
| Dagan | 872 | 488019 | 5775 | 596618 | 23.1% | 8.2% | 0.18 [0.17; 0.20] |
| Daniel | 112 | 6144 | 234 | 8969 | 0.0% | 0.0% | 0.70 [0.56; 0.87] |
| Hall | 66 | 13716 | 902 | 1405 | 0.0% | 0.0% | 0.01 [0.01; 0.01] |
| Lopez Bernal A70 | 587 | 10544 | 15287 | 51955 | 18.3% | 8.2% | 0.19 [0.17; 0.20] |
| Lopez Bernal B70 | 672 | 11052 | 15287 | 51955 | 21.0% | 8.2% | 0.21 [0.19; 0.22] |
| Lopez Bernal A80 | 650 | 4378 | 8988 | 24706 | 22.0% | 8.2% | 0.41 [0.38; 0.44] |
| Pawloski | 193 | 31069 | 1298 | 31069 | 5.2% | 8.0% | 0.15 [0.13; 0.17] |
| Tenforde | 18 | 62 | 146 | 287 | 0.7% | 6.6% | 0.57 [0.38; 0.86] |
| Shotri A | 161 | 2861 | 723 | 9160 | 0.0% | 0.0% | 0.71 [0.60; 0.84] |
| Shotri B | 207 | 5931 | 723 | 9160 | 5.1% | 8.0% | 0.44 [0.38; 0.51] |
| Pritchard | 417 | 373402 | 11697 | 373402 | 0.0% | 0.0% | 0.04 [0.03; 0.04] |
| Swift | 98 | 3210 | 997 | 23931 | 0.0% | 0.0% | 0.73 [0.60; 0.90] |
| Angel | 37 | 5761 | 8 | 741 | 0.2% | 4.3% | 0.59 [0.28; 1.27] |
| Abu-Raddad A | 892 | 2133 | 18075 | 35801 | 0.0% | 0.0% | 0.83 [0.79; 0.87] |
| Abu-Raddad B | 1329 | 2909 | 20177 | 40103 | 0.0% | 0.0% | 0.91 [0.87; 0.95] |
| Jones A | 8 | 2010 | 56 | 3314 | 0.2% | 4.4% | 0.24 [0.11; 0.49] |
| Jones B | 5 | 4867 | 113 | 13970 | 0.1% | 3.7% | 0.13 [0.05; 0.31] |
| Fabiani | 9 | 138 | 169 | 921 | 0.3% | 5.0% | 0.36 [0.19; 0.68] |
| Gras Valenti | 39 | 138 | 31 | 60 | 0.9% | 6.8% | 0.55 [0.38; 0.79] |
| **Total (fixed effect, 95% CI)** | | **981231** | | **1285761** | **100.0%** | **--** | **0.24 [0.23; 0.25]** |
| **Total (random effects, 95% CI)** | | | | | **--** | **100.0%** | **0.27 [0.21; 0.34]** |

Heterogeneity: $Tau^2$ = 0.1657; $Chi^2$ = 459.09, df = 14 ($P < 0.01$); $I^2$ = 97%

**Fig.7a) Influence analysis.** Partially vaccinated, any positive PCR. Influence analysis studied the extreme values in the graphs through different influence measures. Each subplot graphs included the influence estimates for each study of our meta-analysis. Arbitrary cut-offs define an influential case, which is displayed in red.[21] "Hall" resulted as the most influential in partially vaccinated (any positive PCR) meta-analysis.

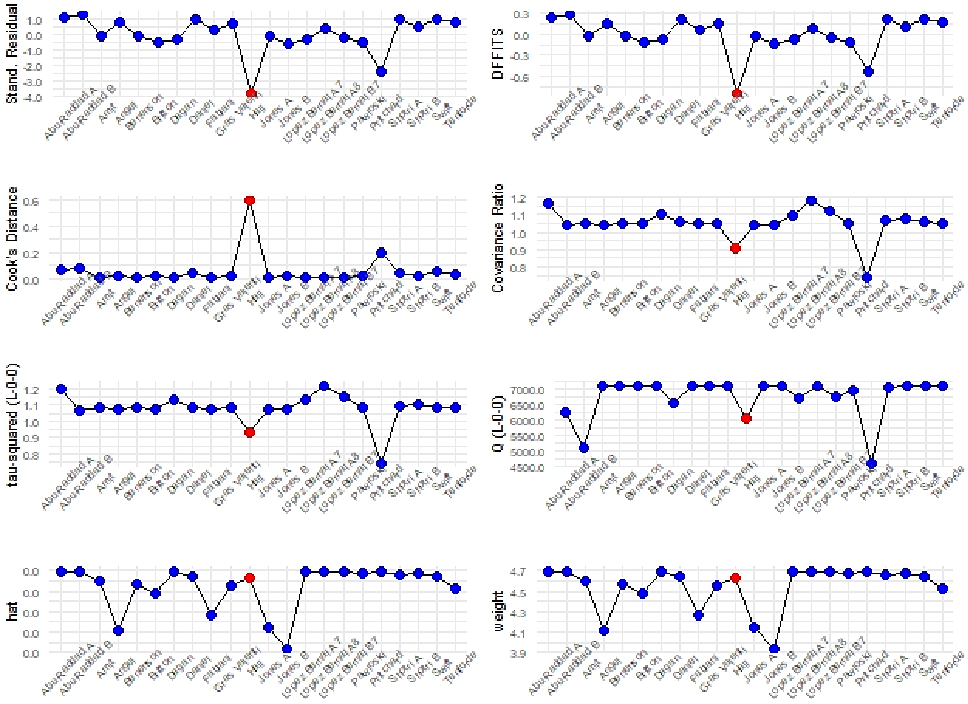

**Fig.7b) Baujat plot.** Partially vaccinated, any positive PCR. Contribute to overall heterogeneity Q (x) and effect size(y). “Pritchard”, “Abu Raddad B” and “Hall” were the most influential on the overall heterogeneity. “Abu Raddad B” contributed the most to the pooled result.

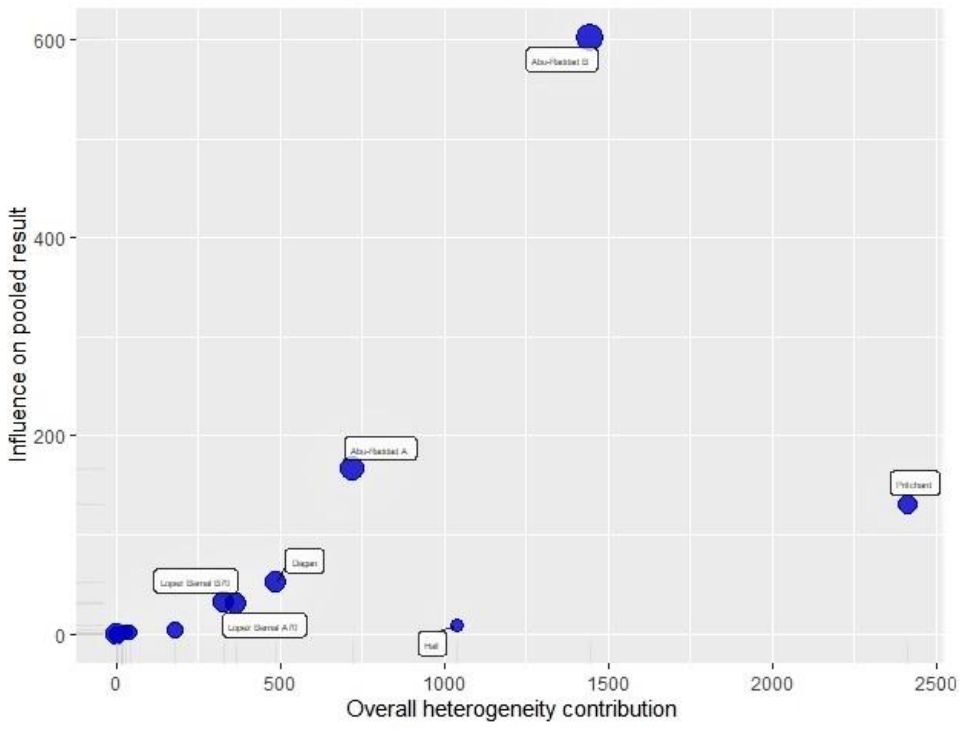


**Fig.8) Leave-One-Out Analysis.** Partially vaccinated, any positive PCR. The first plot shows that by omitting “Abu-Raddad B” study, the I2 is slightly reduced. Moreover, “Swift”, “Daniel” and “Britton” contributed to the overall heterogeneity. In the second plot the overall effect estimate changed with two studies removed. In particular, by removing “Pritchard” the RR increased to 0.29, while RR increased to 0.32 after removing “Hall”.

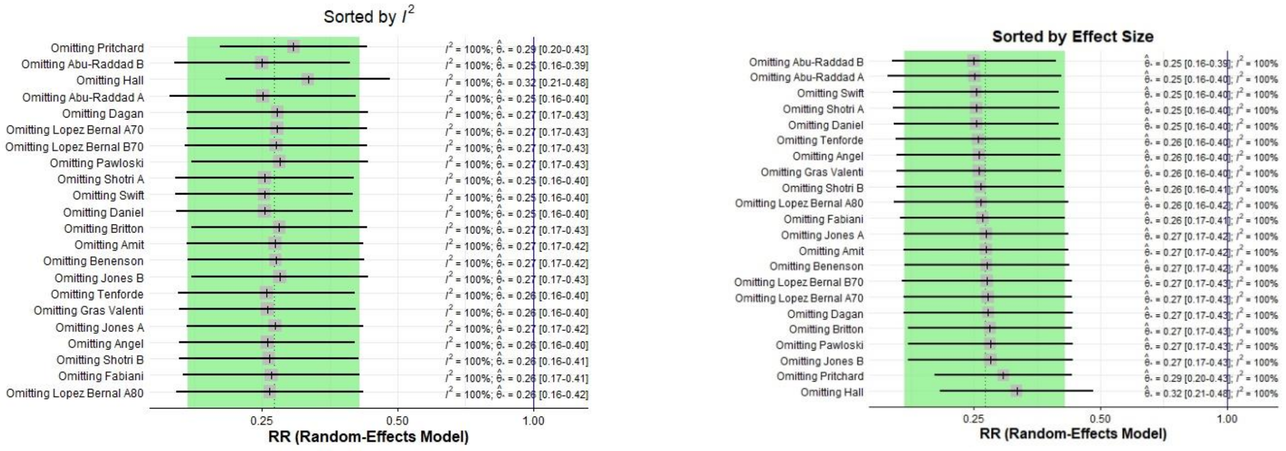

**Fig.9) GOSH Diagnostics.** Partially vaccinated, any positive PCR. GOSH plots did not show clear patterns in our data. The effect size distribution was homogeneous, in contrast, the heterogeneity curve presented an asymmetric distribution with a sharp peak. The three algorithms, k -means, DBSCAN and the Gaussian Mixture Model, detected up to eleven clusters which might potentially contribute to the pooled imbalance, and all of them identified the study “Hall”(6) as contributing to the overall heterogeneity.

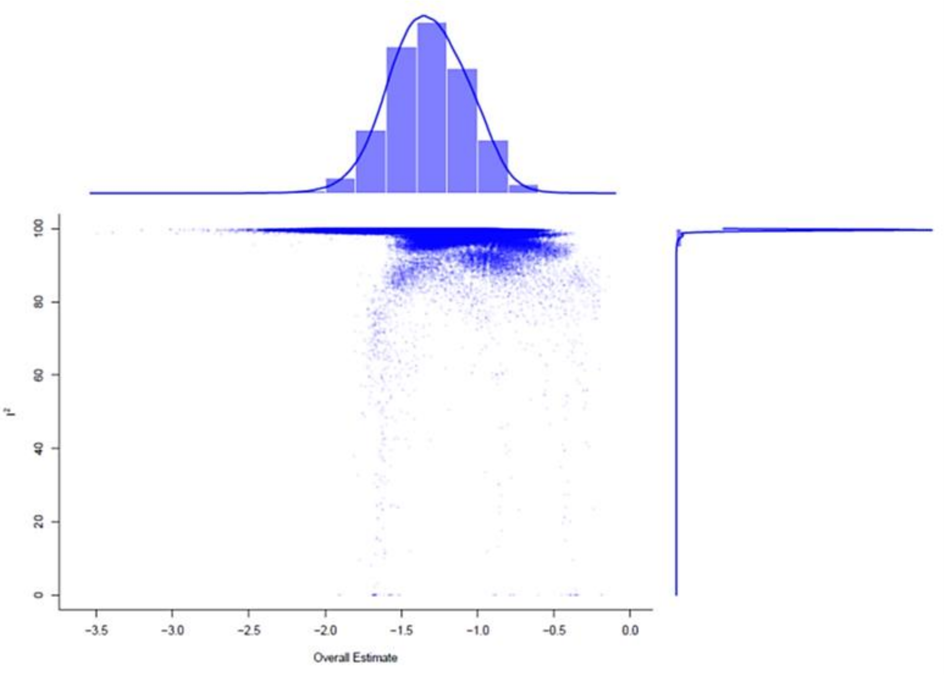


| GOSH Diagnostics |
| --- |
| - Number of K-means clusters detected: 3 |
| - Number of DBSCAN clusters detected: 11 |
| - Number of GMM clusters detected: 9 |
| **Identification of potential outliers** |
| K-means: Study 6 |
| DBSCAN: Study 6, Study 14, Study 11, Study 9, Study 10, Study 21, Study 1, Study 8 |
| Gaussian Mixture Model: Study 6, Study 14, Study 11, Study 9, Study 10, Study 21, Study 1, Study 8 |

#### *A3.2 Fully vaccinated, any Sars-Cov2 positive PCR*

**Table 6)** Outliers, fully vaccinated. The outliers function in full vaccination protocol meta-analysis detected seven outliers.

| **Identified outliers (random-effects model)** | | | | | | |
|---|---|---|---|---|---|---|
| "Dagan", "Hall", "Pawloski", "Pritchard", "Swift", "Abu Raddad A", "Abu Raddad B" | | | | | | |
| **Results with outliers removed** | | | | | | |
| | **RR [95%-CI %]** | | | **W(fixed) %** | **W(random)** | **excluded** |
| **Benenson** | 0.0504 | [0.0259; | 0.0981] | 5.2 | 10.9 | |
| **Britton** | 0.0514 | [0.0238; | 0.1108] | 3.9 | 10.3 | |
| **Dagan** | 0.0136 | [0.0104; | 0.0177] | 0.0 | 0.0 | * |
| **Daniel** | 0.0189 | [0.0070; | 0.0507] | 2.4 | 8.9 | |
| **Hall** | 0.0078 | [0.0039; | 0.0155] | 0.0 | 0.0 | * |
| **Lopez Bernal B80** | 0.1614 | [0.1283; | 0.2031] | 44.2 | 13.0 | |
| **Pawloski** | 0.3638 | [0.3037; | 0.4360] | 0.0 | 0.0 | * |
| **Bouton** | 0.0304 | [0.0187; | 0.0494] | 9.9 | 11.9 | |
| **Tenforde** | 0.1035 | [0.0153; | 0.6994] | 0.6 | 4.7 | |
| **Pritchard** | 0.0062 | [0.0049; | 0.0078] | 0.0 | 0.0 | * |
| **Sansone** | 0.1504 | [0.1104; | 0.2049] | 24.4 | 12.7 | |
| **Swift** | 0.0164 | [0.0114; | 0.0235] | 0.0 | 0.0 | * |
| **Angel** | 0.1448 | [0.0756; | 0.2773] | 5.5 | 11.0 | |
| **Abu Raddad A** | 0.1917 | [0.1473; | 0.2495] | 0.0 | 0.0 | * |
| **Abu Raddad B** | 0.4027 | [0.3533; | 0.4592] | 0.0 | 0.0 | * |
| **Corchado** | 0.2338 | [0.0745; | 0.7337] | 1.8 | 8.0 | |
| **Fabiani** | 0.0316 | [0.0110; | 0.0903] | 2.1 | 8.6 | |
| **Meta-analysis with outliers removed** | | | | | | |
| Number of studies combined: k = 10 | | | | | | |
| | **RR [95%-CI %]** | | | **z** | **p-value** | |
| Fixed effect model | 0.1109 | [0.0952; | 0.1291] | -28.26 | < 0.0001 | |
| Random effects model | 0.0735 | [0.0440; | 0.1228] | -9.97 | < 0.0001 | |
| **Quantifying heterogeneity:** | | | | | | |
| tau^2 = 0.5132 [0.1894; 2.4139]; tau = 0.7164 [0.4352; 1.5537] | | | | | | |
| I^2 = 87.3% [78.6%; 92.4%]; H = 2.80 [2.16; 3.63] | | | | | | |
| Test of heterogeneity: | | | | | | |
| | **Q** | **d.f.** | **p-value** | | | |
| | 70.61 | 9 | <0.0001 | | | |
| **Details on meta-analytical method:** | | | | | | |
| - Inverse variance method | | | | | | |
| - DerSimonian-Laird estimator for tau^2 | | | | | | |
| - Jackson method for confidence interval of tau^2 and tau | | | | | | |

**Fig.10) Forest plot.** Fully vaccinated, any positive PCR test RR ≥7 days after full vaccination schedule fulfilment. Outliers exluded, RE, IV method. Identified outliers: "Dagan", "Hall", "Pawloski", "Pritchard", "Swift", "Abu Raddad A", "Abu Raddad B". The RE reduced meta-analysis showed an overall RR=0.07 ($p<0.0001$), therefore, the RRR of any positive PCR approached 93% one week after the full vaccination protocol. The $I^2$ heterogeneity shrunk (I2=87.3% τ2= 0.5132 [0.1894; 2.4139]), but the heterogeneity test was still significant (Q=70.61, $p<0.0001$).

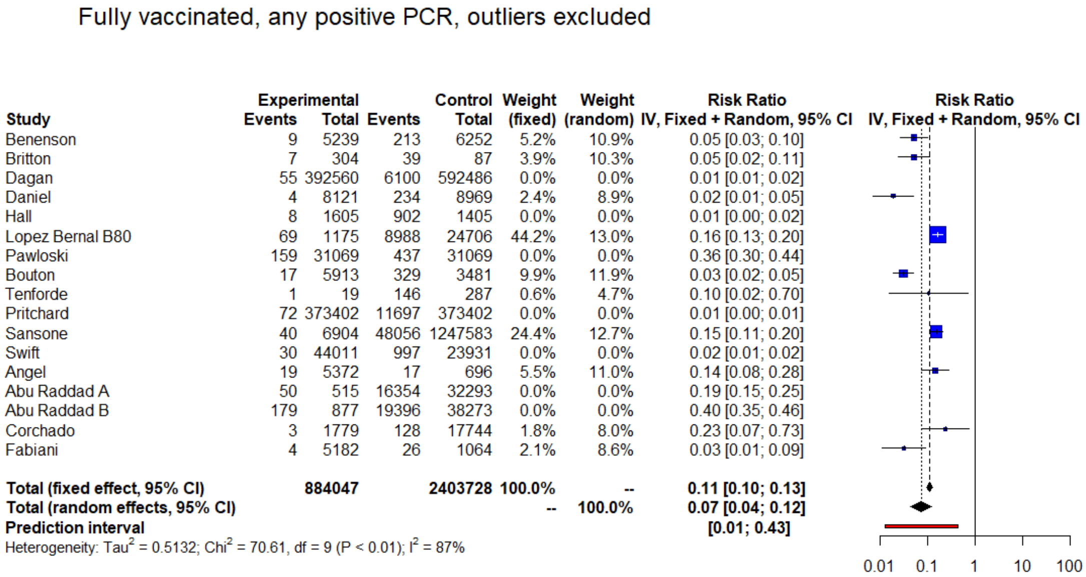

Fully vaccinated, any positive PCR, outliers excluded

| Study | Experimental Events | Experimental Total | Control Events | Control Total | Weight (fixed) | Weight (random) | Risk Ratio IV, Fixed + Random, 95% CI |
|---|---|---|---|---|---|---|---|
| Benenson | 9 | 5239 | 213 | 6252 | 5.2% | 10.9% | 0.05 [0.03; 0.10] |
| Britton | 7 | 304 | 39 | 87 | 3.9% | 10.3% | 0.05 [0.02; 0.11] |
| Dagan | 55 | 392560 | 6100 | 592486 | 0.0% | 0.0% | 0.01 [0.01; 0.02] |
| Daniel | 4 | 8121 | 234 | 8969 | 2.4% | 8.9% | 0.02 [0.01; 0.05] |
| Hall | 8 | 1605 | 902 | 1405 | 0.0% | 0.0% | 0.01 [0.00; 0.02] |
| Lopez Bernal B80 | 69 | 1175 | 8988 | 24706 | 44.2% | 13.0% | 0.16 [0.13; 0.20] |
| Pawloski | 159 | 31069 | 437 | 31069 | 0.0% | 0.0% | 0.36 [0.30; 0.44] |
| Bouton | 17 | 5913 | 329 | 3481 | 9.9% | 11.9% | 0.03 [0.02; 0.05] |
| Tenforde | 1 | 19 | 146 | 287 | 0.6% | 4.7% | 0.10 [0.02; 0.70] |
| Pritchard | 72 | 373402 | 11697 | 373402 | 0.0% | 0.0% | 0.01 [0.00; 0.01] |
| Sansone | 40 | 6904 | 48056 | 1247583 | 24.4% | 12.7% | 0.15 [0.11; 0.20] |
| Swift | 30 | 44011 | 997 | 23931 | 0.0% | 0.0% | 0.02 [0.01; 0.02] |
| Angel | 19 | 5372 | 17 | 696 | 5.5% | 11.0% | 0.14 [0.08; 0.28] |
| Abu Raddad A | 50 | 515 | 16354 | 32293 | 0.0% | 0.0% | 0.19 [0.15; 0.25] |
| Abu Raddad B | 179 | 877 | 19396 | 38273 | 0.0% | 0.0% | 0.40 [0.35; 0.46] |
| Corchado | 3 | 1779 | 128 | 17744 | 1.8% | 8.0% | 0.23 [0.07; 0.73] |
| Fabiani | 4 | 5182 | 26 | 1064 | 2.1% | 8.6% | 0.03 [0.01; 0.09] |
| **Total (fixed effect, 95% CI)** | | **884047** | | **2403728** | **100.0%** | -- | **0.11 [0.10; 0.13]** |
| **Total (random effects, 95% CI)** | | | | | -- | **100.0%** | **0.07 [0.04; 0.12]** |
| **Prediction interval** | | | | | | | **[0.01; 0.43]** |

Heterogeneity: $Tau^2 = 0.5132$; $Chi^2 = 70.61$, df = 9 ($P < 0.01$); $I^2 = 87\%$



**Fig. 11a) Influence analysis.** Fully vaccinated, any positive PCR. Influential analysis subplot graphs included the influence estimates for each study in fully vaccinated meta-analysis. No study exceeded the cut-offs to define an influential case; however, "Pritchard" study needed further investigation.[21]

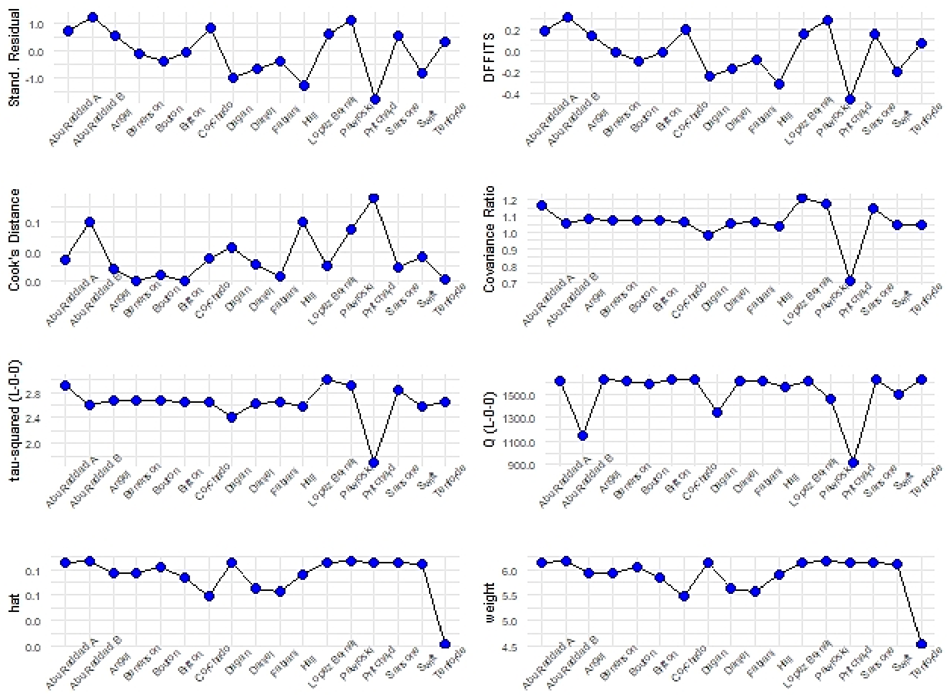

**Fig. 11b) Baujat plot.** Fully vaccinated, any positive PCR. Contribute to overall heterogeneity (Q) and effect size. “Pritchard” and “Abu-Raddab B” contributed to the pooled imbalance. “Abu Raddad B” was the most influential on pooled effect size.

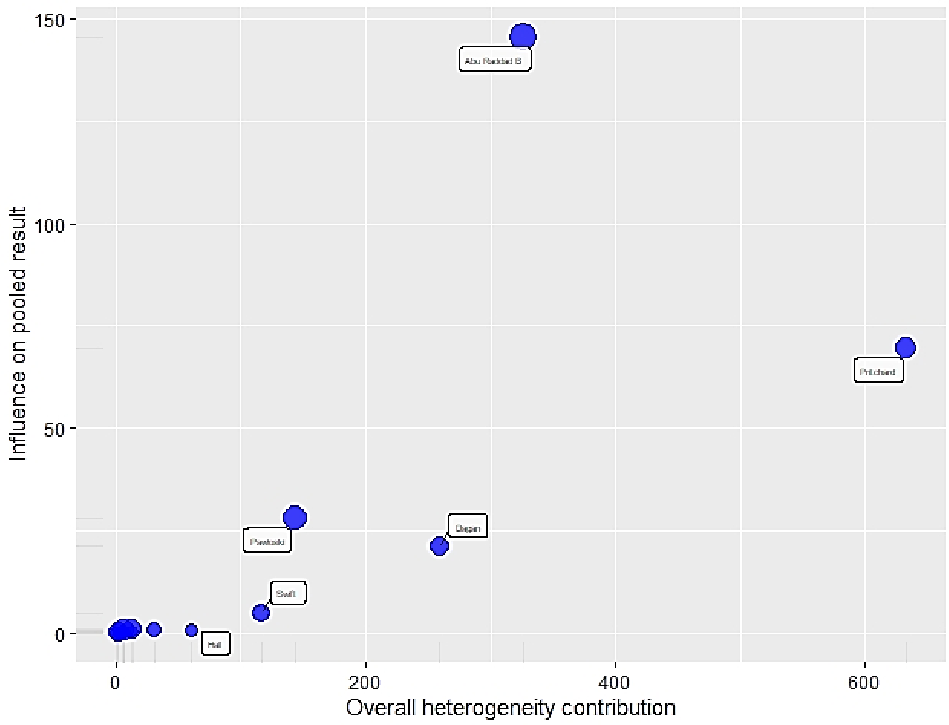


**Fig.12) Leave one out analysis**. Fully vaccinated, any positive PCR. The $I^2$ heterogeneity, printed in the first plot, slightly diminished after omitting “Pritchard”, “Abu Raddad B”, “Pawloski” and “Corchado”. The first plot indicated how the omission of each study influenced the overall effect size estimate. Moreover, “Abu Raddad B”or “Pawloski” or “Corchado” omission reduced the RR and the upper 95%CI, while RR rose to 0.07 after removing “Hall” or “Pritchard”.

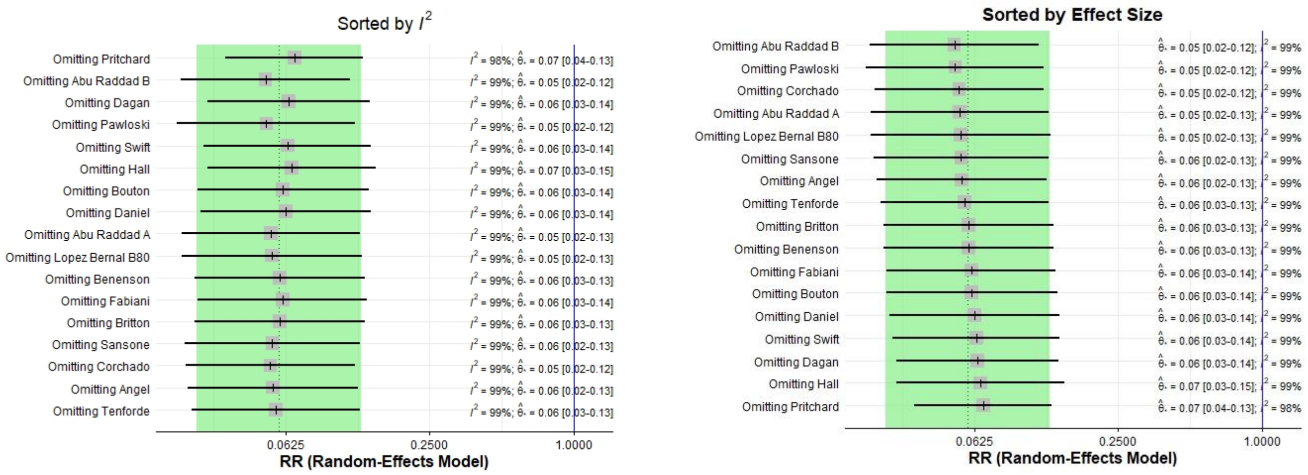

**Fig.13) GOSH Diagnostics.** Fully vaccinated, any positive PCR. GOSH plots did not show any pattern. The effect size distribution is symmetric, in contrast, the heterogeneity curve ($I^2$) is right skewed and a sharp peak. The three algorithms, k -means, DBSCAN and the Gaussian Mixture Model, detected up to 15 entries which might potentially contribute to the pooled imbalance, and all of them identified "Pritchard"(10) as contributing to the overall heterogeneity.

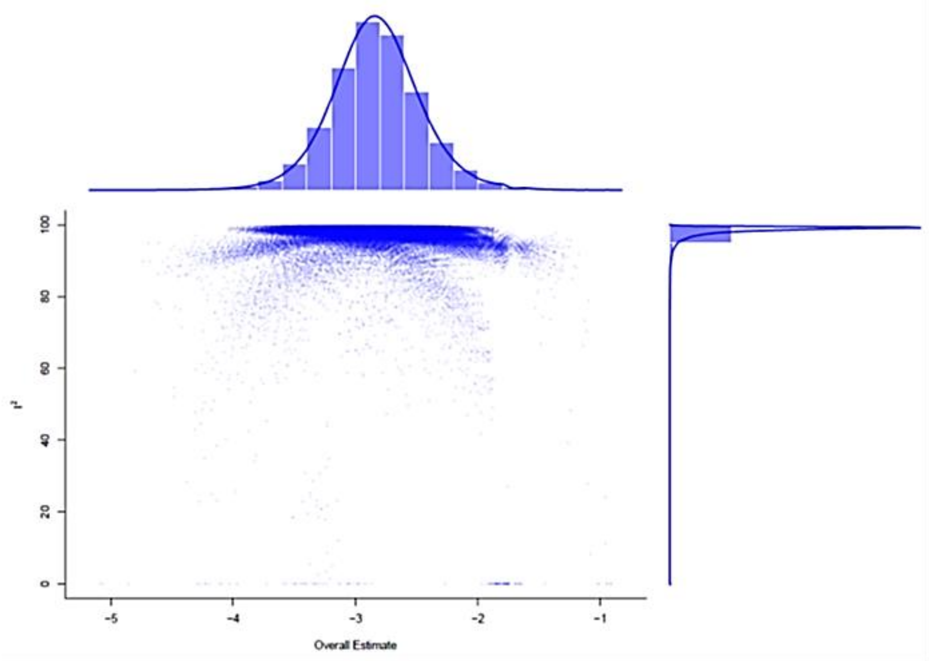


| GOSH Diagnostics |
| --- |
| - Number of K-means clusters detected: 3 |
| - Number of DBSCAN clusters detected: 15 |
| - Number of GMM clusters detected: 9 |
| **Identification of potential outliers** |
| - K-means: Study 10, Study 9 |
| - DBSCAN: Study 15, Study 7, Study 14, Study 5, Study 13, Study 3, Study 10 |
| - Gaussian Mixture Model: Study 15, Study 7, Study 14, Study 5, Study 13, Study 3, Study 10 |

#### *A.3.3 At least one dose effectiveness, any Sars-Cov2 positive PCR*

**Table 7)** Outliers, at least one dose, any positive PCR. In meta-analysis of individuals vaccinated with at least one dose, the outliers function detected nine extreme estimates.

| **Identified outliers (random-effects model)** | | | | | | |
|---|---|---|---|---|---|---|
| "Hall", "Lopez Bernal C80", "Moustsen Helms R", "Pawloski", "Tande", "Bouton", "Monge", "Mason", "Menni A" | | | | | | |
| **Results with outliers removed** | | | | | | |
| | **RR [95%-CI %]** | | | **W(fixed) %** | **W(random)** | **excluded** |
| **Amit** | 0.2391 | [0.1777; | 0.3216] | 1.6 | 10.5 | |
| **Benenson** | 0.2992 | [0.2225; | 0.4025] | 1.6 | 10.5 | |
| **Britton** | 0.1899 | [0.1266; | 0.2847] | 0.9 | 8.7 | |
| **Dagan** | 0.1520 | [0.1419; | 0.1629] | 29.8 | 13.5 | |
| **Hall** | 0.0084 | [0.0067; | 0.0106] | 0.0 | 0.0 | * |
| **Lopez Bernal C70** | 0.2251 | [0.2103; | 0.2410] | 30.6 | 13.5 | |
| **Lopez Bernal C80** | 0.4985 | [0.4671; | 0.5320] | 0.0 | 0.0 | * |
| **Lumley A** | 0.0879 | [0.0649; | 0.1192] | 1.5 | 10.4 | |
| **Lumley B** | 0.0726 | [0.0411; | 0.1283] | 0.4 | 6.4 | |
| **Moustsen Helms H** | 0.1264 | [0.1120; | 0.1427] | 9.7 | 13.1 | |
| **Moustsen Helms R** | 0.0276 | [0.0240; | 0.0318] | 0.0 | 0.0 | * |
| **Pawloski** | 0.3979 | [0.3452; | 0.4586] | 0.0 | 0.0 | * |
| **Tande** | 0.4410 | [0.3252; | 0.5981] | 0.0 | 0.0 | * |
| **Bouton** | 0.0432 | [0.0296; | 0.0630] | 0.0 | 0.0 | * |
| **Monge** | 0.5751 | [0.5623; | 0.5882] | 0.0 | 0.0 | * |
| **Mason** | 0.5407 | [0.4953; | 0.5902] | 0.0 | 0.0 | * |
| **Menni A** | 0.3127 | [0.3006; | 0.3253] | 0.0 | 0.0 | * |
| **Menni B** | 0.1481 | [0.1371; | 0.1600] | 23.8 | 13.5 | |
| **Meta-analysis with outliers removed** | | | | | | |
| Number of studies combined: k = 9 | | | | | | |
| | **RR [95%-CI %]** | | | **z** | **p-value** | |
| Fixed effect model | 0.1687 | [0.1625; | 0.1752] | -92.58 | 0 | |
| Random effects model | 0.1613 | [0.1325; | 0.1964] | -18.19 | < 0.0001 | |
| **Quantifying heterogeneity:** | | | | | | |
| tau^2 = 0.0731 [0.0408; 0.5199]; tau = 0.2704 [0.2020; 0.7210] | | | | | | |
| I^2 = 94.9% [92.2%; 96.6%]; H = 4.42 [3.58; 5.46] | | | | | | |
| Test of heterogeneity: | | | | | | |
| | **Q** | **d.f.** | **p-value** | | | |
| | 156.58 | 8 | <0.0001 | | | |
| **Details on meta-analytical method:** | | | | | | |
| - Inverse variance method | | | | | | |
| - DerSimonian-Laird estimator for tau^2 | | | | | | |
| - Jackson method for confidence interval of tau^2 and tau | | | | | | |

**Fig.14) Forest plot.** At least one dose, any positive PCR RR ≥14 days after first dose. Outliers excluded, RE, IV method. Identified outliers: "Hall", "Lopez Bernal C80", "Moustsen Helms R", "Pawloski", "Tande", "Bouton", "Monge", "Mason", "Menni A". The reduced meta-analysis output showed an overall RR=0.16 with RRR= 84%. Heterogeneity: τ2 = 0.0731 [0.0408; 0.5199]; τ = 0.2704 [0.2020; 0.7210]; I^2 = 94.9% [92.2%; 96.6%]; H = 4.42 [3.58; 5.46]. Test of heterogeneity: Q =156.58 (d.f.= 8; p-value< 0.0001).

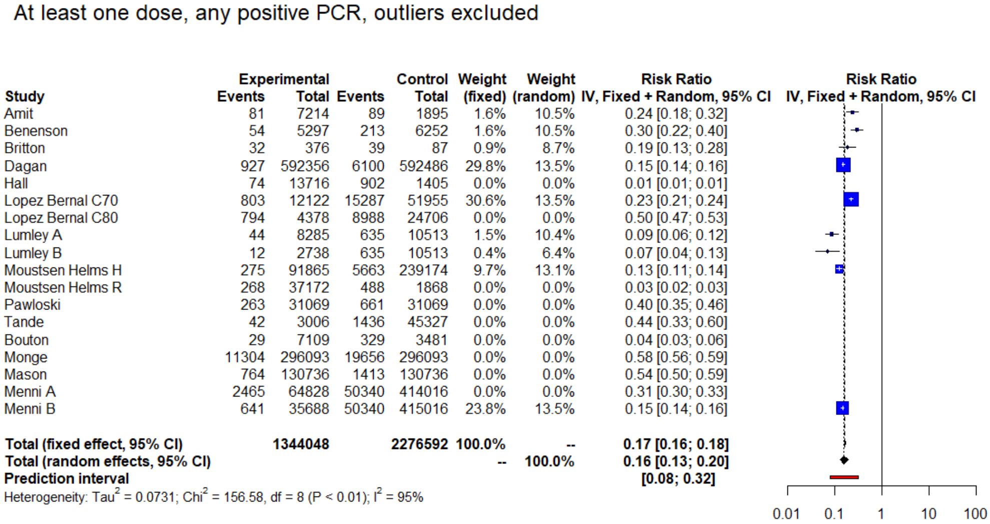


| Study | Experimental Events | Experimental Total | Control Events | Control Total | Weight (fixed) | Weight (random) | Risk Ratio IV, Fixed + Random, 95% CI |
|---|---|---|---|---|---|---|---|
| Amit | 81 | 7214 | 89 | 1895 | 1.6% | 10.5% | 0.24 [0.18; 0.32] |
| Benenson | 54 | 5297 | 213 | 6252 | 1.6% | 10.5% | 0.30 [0.22; 0.40] |
| Britton | 32 | 376 | 39 | 87 | 0.9% | 8.7% | 0.19 [0.13; 0.28] |
| Dagan | 927 | 592356 | 6100 | 592486 | 29.8% | 13.5% | 0.15 [0.14; 0.16] |
| Hall | 74 | 13716 | 902 | 1405 | 0.0% | 0.0% | 0.01 [0.01; 0.01] |
| Lopez Bernal C70 | 803 | 12122 | 15287 | 51955 | 30.6% | 13.5% | 0.23 [0.21; 0.24] |
| Lopez Bernal C80 | 794 | 4378 | 8988 | 24706 | 0.0% | 0.0% | 0.50 [0.47; 0.53] |
| Lumley A | 44 | 8285 | 635 | 10513 | 1.5% | 10.4% | 0.09 [0.06; 0.12] |
| Lumley B | 12 | 2738 | 635 | 10513 | 0.4% | 6.4% | 0.07 [0.04; 0.13] |
| Moustsen Helms H | 275 | 91865 | 5663 | 239174 | 9.7% | 13.1% | 0.13 [0.11; 0.14] |
| Moustsen Helms R | 268 | 37172 | 488 | 1868 | 0.0% | 0.0% | 0.03 [0.02; 0.03] |
| Pawloski | 263 | 31069 | 661 | 31069 | 0.0% | 0.0% | 0.40 [0.35; 0.46] |
| Tande | 42 | 3006 | 1436 | 45327 | 0.0% | 0.0% | 0.44 [0.33; 0.60] |
| Bouton | 29 | 7109 | 329 | 3481 | 0.0% | 0.0% | 0.04 [0.03; 0.06] |
| Monge | 11304 | 296093 | 19656 | 296093 | 0.0% | 0.0% | 0.58 [0.56; 0.59] |
| Mason | 764 | 130736 | 1413 | 130736 | 0.0% | 0.0% | 0.54 [0.50; 0.59] |
| Menni A | 2465 | 64828 | 50340 | 414016 | 0.0% | 0.0% | 0.31 [0.30; 0.33] |
| Menni B | 641 | 35688 | 50340 | 415016 | 23.8% | 13.5% | 0.15 [0.14; 0.16] |
| **Total (fixed effect, 95% CI)** | | **1344048** | | **2276592** | **100.0%** | **--** | **0.17 [0.16; 0.18]** |
| **Total (random effects, 95% CI)** | | | | | **--** | **100.0%** | **0.16 [0.13; 0.20]** |
| **Prediction interval** | | | | | | | **[0.08; 0.32]** |

Heterogeneity: $Tau^2 = 0.0731$; $Chi^2 = 156.58$, df = 8 ($P < 0.01$); $I^2 = 95\%$

**Fig. 15a) Influence analysis.** At least one dose, any positive PCR. “Hall” (red dot) resulted as the most influential study as it exceeded the cut-offs to defined by Viechtbauer and Cheung in each subplot.[21]

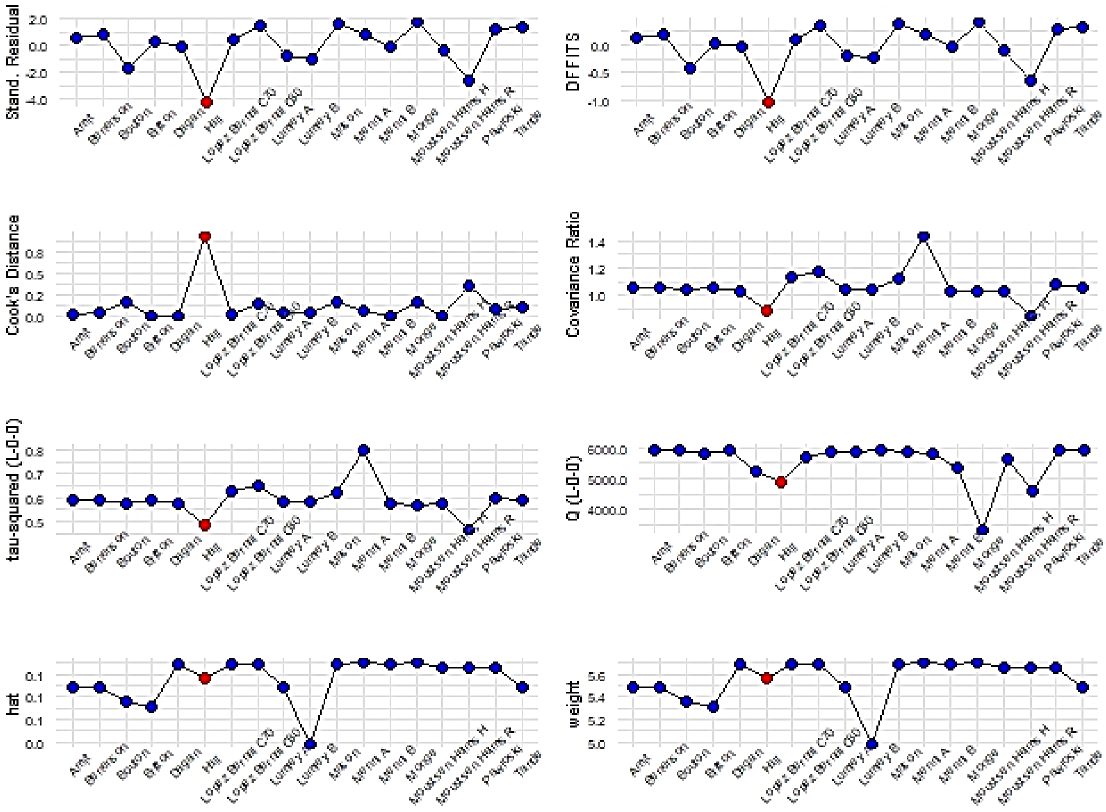

**Fig.15b) Baujat plot.** At least one dose any positive PCR. Contribute to overall heterogeneity, Q (x) and effect size (y). “Hall” and “"Moustsen Helms R" and “Monge” contributed to the pooled imbalance. “Monge” was the most influential on pooled effect size.

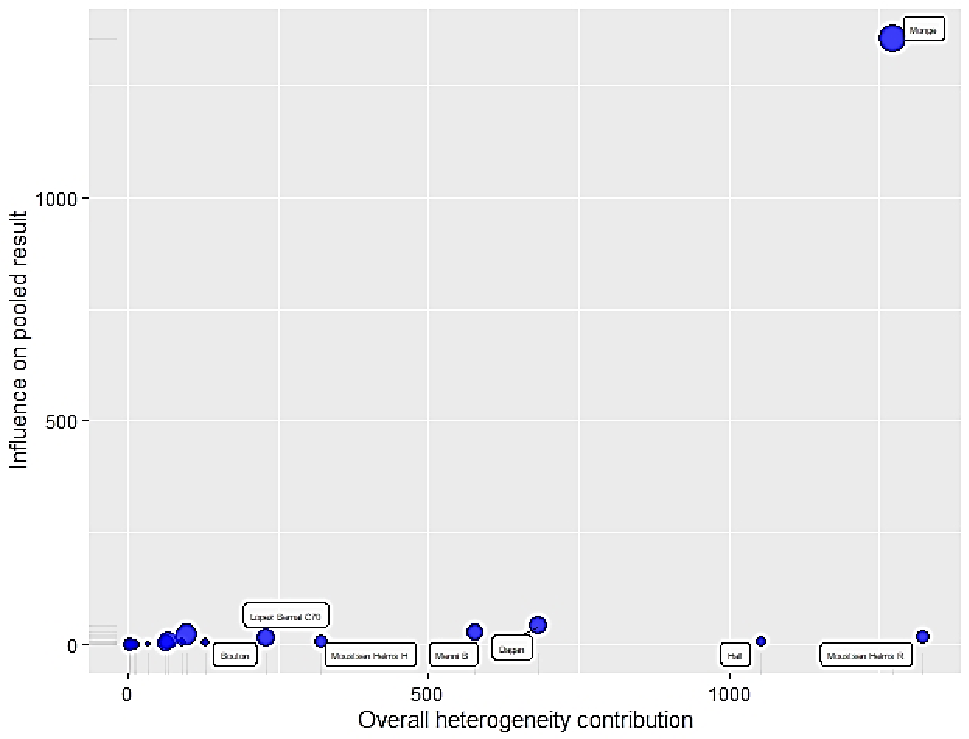


**Fig.16) Leave one out analysis.** At least one dose, any positive PCR. The first plot displayed the change on the I2 heterogeneity attained by removing one study. The second plot indicated how the omission of each study influenced the overall RR estimate. The best result in both cases was achieved by omitting “Monge”.

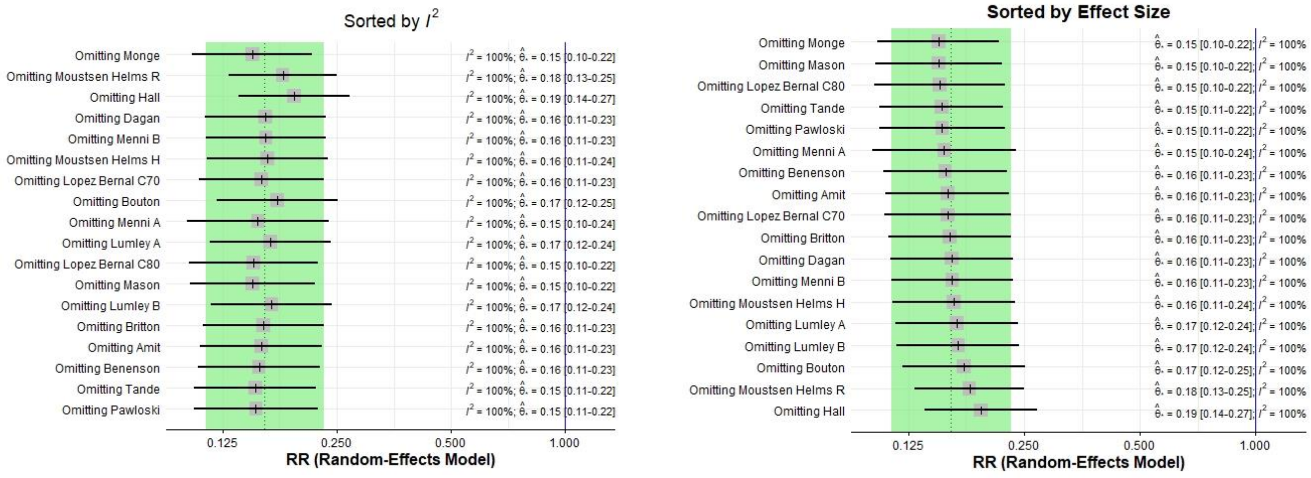

**Fig.17) GOSH Diagnostics, at least one dose, any positive PCR.** GOSH plots did not show any pattern and corroborated the outliers tests and the influence analyses. The effect size distribution was symmetric, while the heterogeneity curve ($I^2$) was right skewed. The three algorithms, k -means, DBSCAN and the Gaussian Mixture Model, detected up to eight entries which might contribute to the pooled imbalance, as well as to the overall heterogeneity, in particular "Hall" (5), "Monge" (15) and "Lopez Bernal C80" on ≥80 years (7).

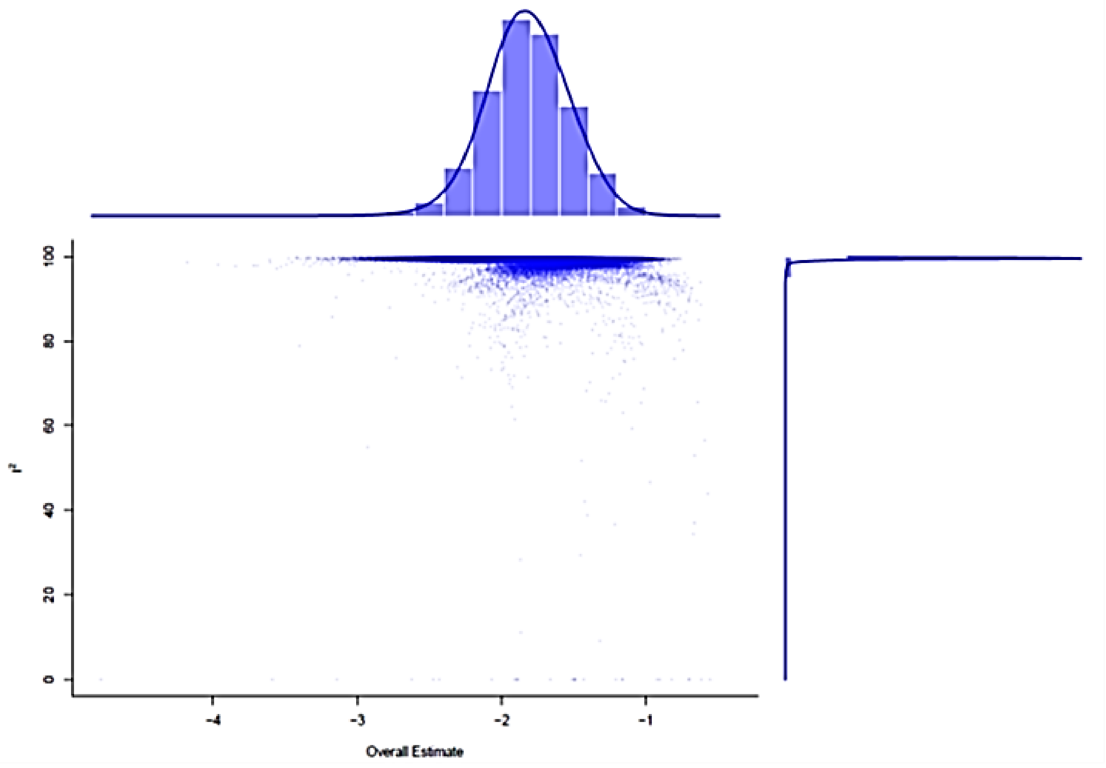


| GOSH Diagnostics |
| --- |
| Number of K-means clusters detected: 3<br>Number of DBSCAN clusters detected: 6<br>Number of GMM clusters detected: 8 |
| **Identification of potential outliers** |
| K-means: Study 111, Study 15, Study 5<br>DBSCAN: Study 5, Study 7<br>Gaussian Mixture Model: Study 5, Study 7 |

#### *A3.4 Sars-Cov2 effectiveness, symptomatic positive PCR*

**Table 8)** Outliers, partially vaccinated, symptomatic Sars-Cov-2 PCR. The outliers function identified "Pritchard".

| | | | | | | |
|---|---|---|---|---|---|---|
| **Identified outliers (random-effects model)** | | | | | | |
| "Pritchard" | | | | | | |
| **Results with outliers removed** | | | | | | |
| | **RR [95%-CI %]** | | | **W(fixed) %** | **W(random)** | **excluded** |
| **Amit** | 0.0482 | [0.0254; | 0.0914] | 0.6 | 12.0 | |
| **Dagan** | 0.6899 | [0.6549; | 0.7267] | 91.6 | 14.5 | |
| **Hyams A** | 0.4217 | [0.2703; | 0.6577] | 1.3 | 13.2 | |
| **Hyams B** | 0.3441 | [0.1763; | 0.6715] | 0.6 | 11.9 | |
| **Pritchard** | 0.0270 | [0.0231; | 0.0317] | 0.0 | 0.0 | * |
| **Swift** | 0.5552 | [0.4336; | 0.7111] | 4.1 | 14.1 | |
| **Angel** | 0.1078 | [0.0673; | 0.1726] | 1.1 | 13.1 | |
| **Jones** | 0.2515 | [0.0879; | 0.7197] | 0.2 | 9.4 | |
| **Fabiani** | 0.5753 | [0.2985; | 1.1086] | 0.6 | 11.9 | |
| **Meta-analysis with outliers removed** | | | | | | |
| Number of studies combined: k =8 | | | | | | |
| | **RR [95%-CI %]** | | | **z** | **p-value** | |
| Fixed effect model | 0.6503 | [0,6187; | 0.6836] | -16.93 | < 0.0001 | |
| Random effects model | 0.2928 | [0.1704; | 0.5029] | -4.45 | < 0.0001 | |
| **Quantifying heterogeneity:** | | | | | | |
| tau^2 = 0.5259 [0.2362; 4.0041]; tau = 0.7252 [0.4860; 2.0010] | | | | | | |
| I^2 = 94.9% [91.9%; 96.7%]; H = 4.41 [3.52; 5.53] | | | | | | |
| Test of heterogeneity: | | | | | | |
| | **Q** | **d.f.** | **p-value** | | | |
| | 136.23 | 7 | <0.0001 | | | |
| **Details on meta-analytical method:** | | | | | | |
| - Inverse variance method | | | | | | |
| - DerSimonian-Laird estimator for tau^2 | | | | | | |
| - Jackson method for confidence interval of tau^2 and tau | | | | | | |

**Fig.18) Forest plot.** Partially vaccinated, symptomatic Covid-19 PCR RR ≥14 days after first dose. Outliers excluded, RE, IV method. The RE reduced meta-analysis was significant ($p< 0.0001$) with an overall RR=0.29 and RRR= 71%.Heterogeneity: $\tau2$=0.5259[0.2362; 4.0041]; $\tau$= 0.7252 [0.4860; 2.0010]; I2 = 94.9% [91.9%; 96.7%]; H = 4.41 [3.52; 5.53]. Test of heterogeneity: Q=136.23 (d.f.=7; p-value< 0.0001).

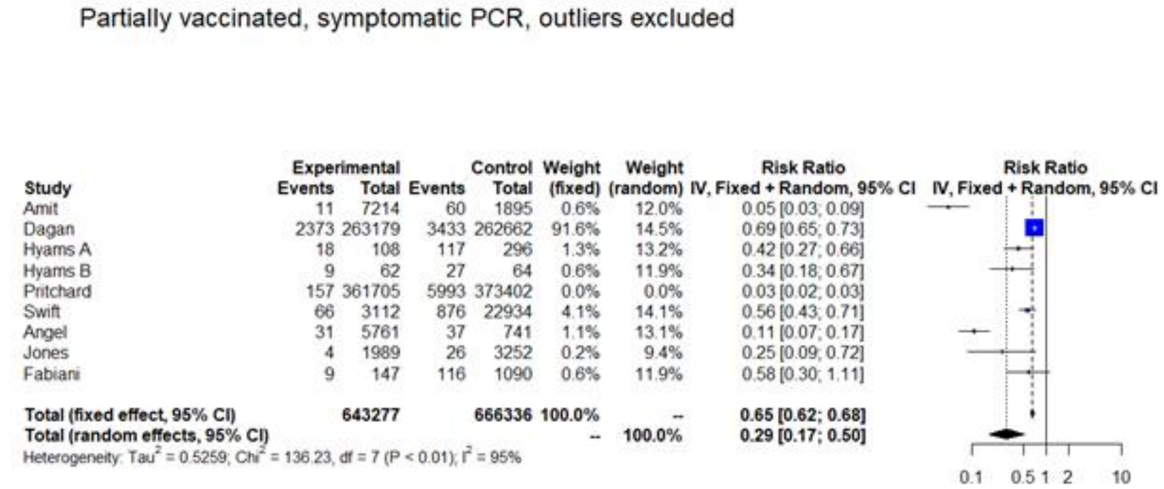


**Fig.19a) Influence analysis.** Partially vaccinated, symptomatic positive Sars-Cov-2 PCR. “Pritchard” (red dot) results as the most influential study as it exceeds the cut-offs defined by Viechtbauerand Cheung in any subplot. [21]

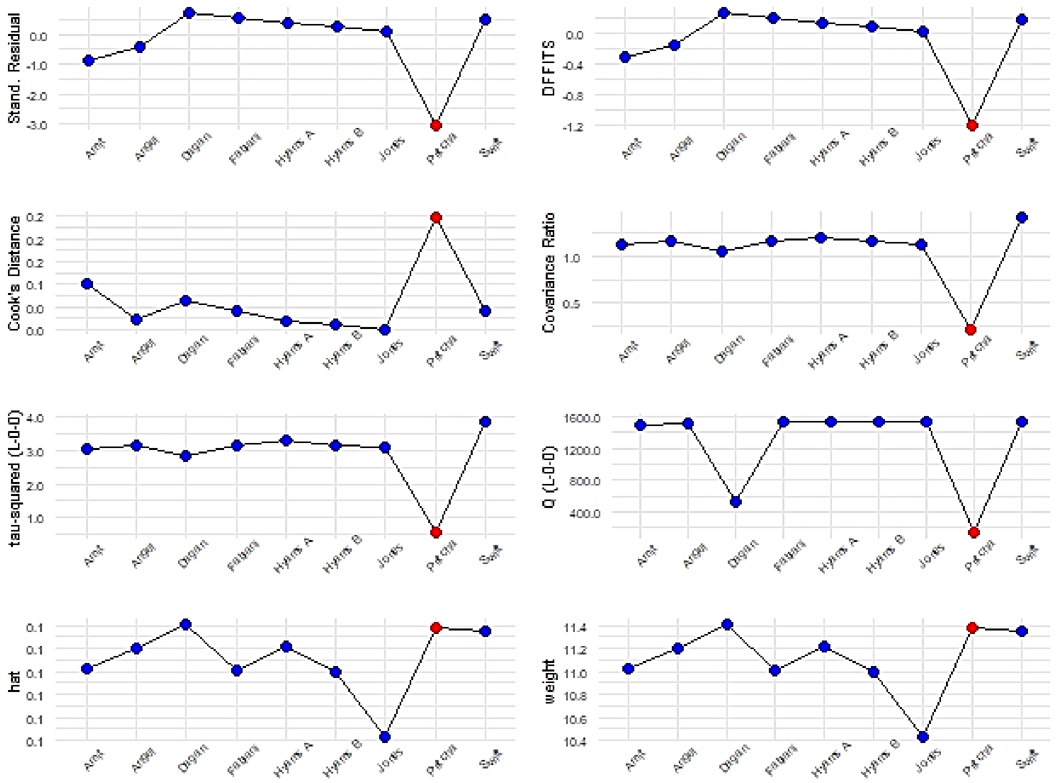

**Fig.19b) Baujat plot.** Partially vaccinated, symptomatic PCR. Contribute to overall heterogeneity, Q (x) and effect size (y)**.** “Pritchard” study contributed to the pooled imbalance. “Dagan” was the most influential on pooled effect size.

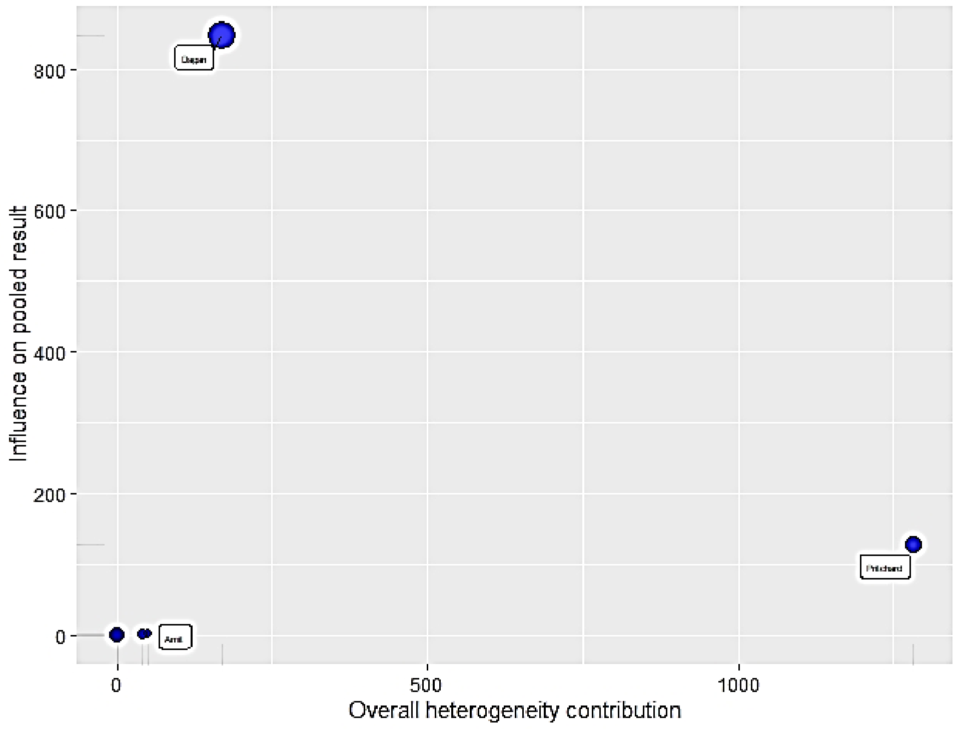


**Fig.20) Leave one out analysis.** Partially vaccinated, symptomatic PCR. The first plot displays the change on the I2 heterogeneity attained by removing one study at each step. The best result on the pooled imbalance was achieved by omitting “Pritchard”. The second plot indicates how the omission of “Dagan” influenced the overall RR estimate.

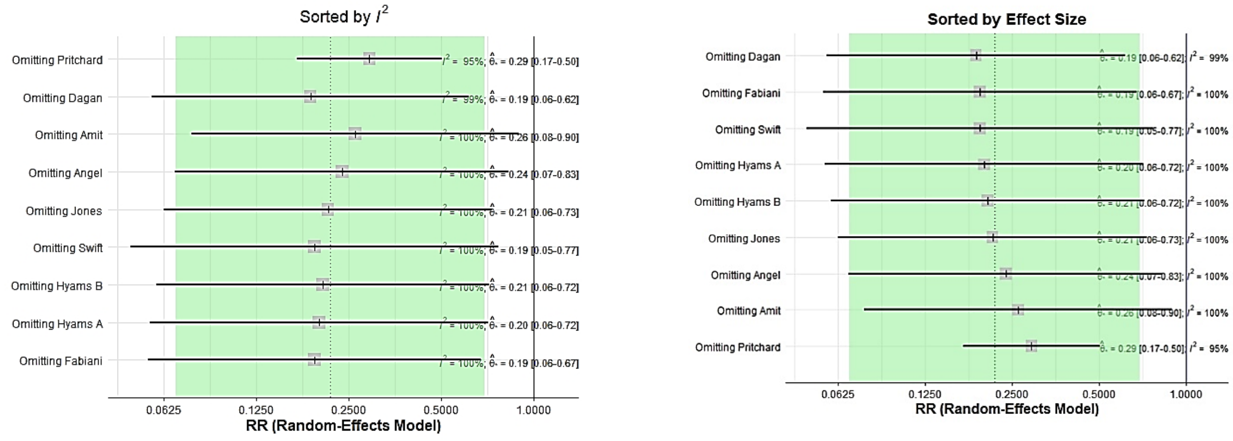

**Fig.21) GOSH Diagnostics.** Partially vaccinated, symptomatic PCR. GOSH plots did not show any pattern, but a fatter right tail on the effect size distribution that was ultimately symmetric. The heterogeneity curve (I2) was right skewed with a sharp peak. The three algorithms, k -means, DBSCAN and the Gaussian Mixture Model, detected up to six clusters which might potentially contribute to the overall heterogeneity, including "Pritchard"(5).

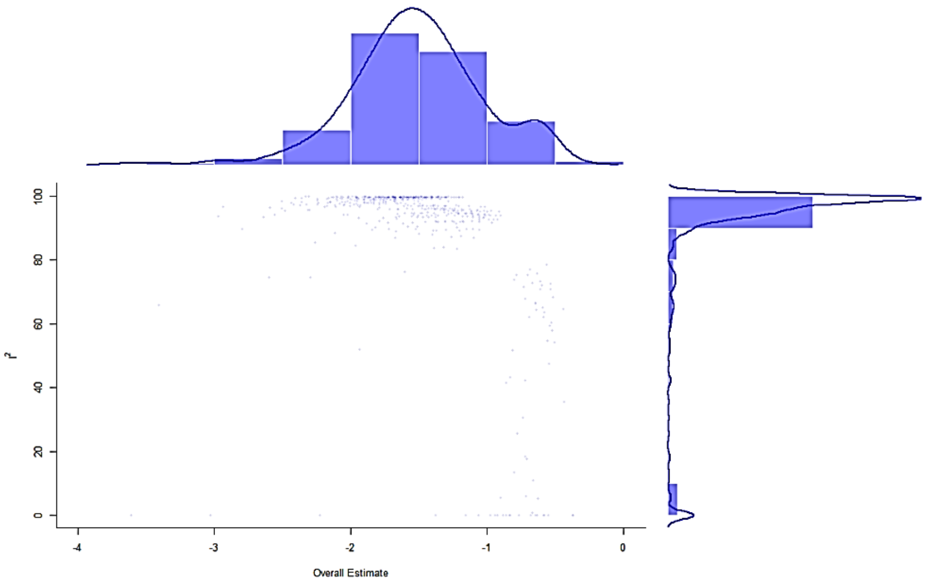


| GOSH Diagnostics |
|---|
| - Number of K-means clusters detected: 3 |
| - Number of DBSCAN clusters detected: 5 |
| - Number of GMM clusters detected: 6 |
| **Identification of potential outliers** |
| - K-means: Study 5 |
| - DBSCAN: Study 3, Study 2 |
| - Gaussian Mixture Model: Study 3, Study 2 |

**Tab.9)** Outliers fully vaccinated, symptomatic Sars-Cov-2 PCR. The outliers function identified four entries in fully vaccinated meta-analysis: "Pritchard", "Swift", "Abu-Raddad B", "Abu-Raddad C".

| **Identified outliers (random-effects model)** | | | | | | |
|---|---|---|---|---|---|---|
| "Pritchard", "Swift", "Abu-Raddad B", "Abu-Raddad C" | | | | | | |
| **Results with outliers removed** | | | | | | |
| | **RR [95%-CI %]** | | | **W(fixed) %** | **W(random)** | **excluded** |
| **Dagan** | 0.0911 | [0.0546; | 0.1520] | 20.3 | 27.9 | |
| **Pritchard** | 0.0021 | [0.0012; | 0.0036] | 0.0 | 0.0 | * |
| **Swift** | 0.0131 | [0.0086; | 0.0200] | 0.0 | 0.0 | * |
| **Angel** | 0.0273 | [0.0128; | 0.0582] | 9.2 | 24.4 | |
| **Abu Raddad A** | 0.1678 | [0.1266; | 0.2223] | 67.1 | 30.4 | |
| **Abu Raddad B** | 0.3598 | [0.3111; | 0.4162] | 0.0 | 0.0 | * |
| **Abu Raddad C** | 0.4001 | [0.3481; | 0.4599] | 0.0 | 0.0 | * |
| **Fabiani** | 0.1401 | [0.0396; | 0.4957] | 3.3 | 17.3 | |
| **Meta-analysis with outliers removed** | | | | | | |
| Number of studies combined: k =8 | | | | | | |
| | **RR [95%-CI %]** | | | **z** | **p-value** | |
| Fixed effect model | 0.1246 | [0,0989; | 0.1569] | -17.70 | < 0.0001 | |
| Random effects model | 0.0881 | [0.0402; | 0.1927] | -6.08 | < 0.0001 | |
| **Quantifying heterogeneity:** | | | | | | |
| tau^2 = 0.5049 [0.0877; 9.5307]; tau = 0.7105 [0.2961; 3.0872] | | | | | | |
| I^2 = 85.8% [65.3%; 94.2%]; H = 2.66 [1.70; 4.16] | | | | | | |
| Test of heterogeneity: | | | | | | |
| | **Q** | **d.f.** | **p-value** | | | |
| | 21.18 | 3 | <0.0001 | | | |
| **Details on meta-analytical method:** | | | | | | |
| - Inverse variance method | | | | | | |
| - DerSimonian-Laird estimator for tau^2 | | | | | | |
| - Jackson method for confidence interval of tau^2 and tau | | | | | | |

**Fig.22) Forest plot.** Fully vaccinated, symptomatic Covid-19 PCR RR ≥7 days after second dose. Outliers excluded, RE, IV method. Identified outliers: "Pritchard", "Swift", "Abu-Raddad B", "Abu Raddad C". Overall RR=0.09, RRR=91%. Heterogeneity: τ2 = 0.5049 [0.0877; 9.5307]; τ= 0.7105 [0.2961; 3.0872]; I2= 85.8% [65.3%; 94.2%]; H = 2.66 [1.70; 4.16]. Test of heterogeneity: Q= 21.18 (d.f.=3; p-value< 0.0001).

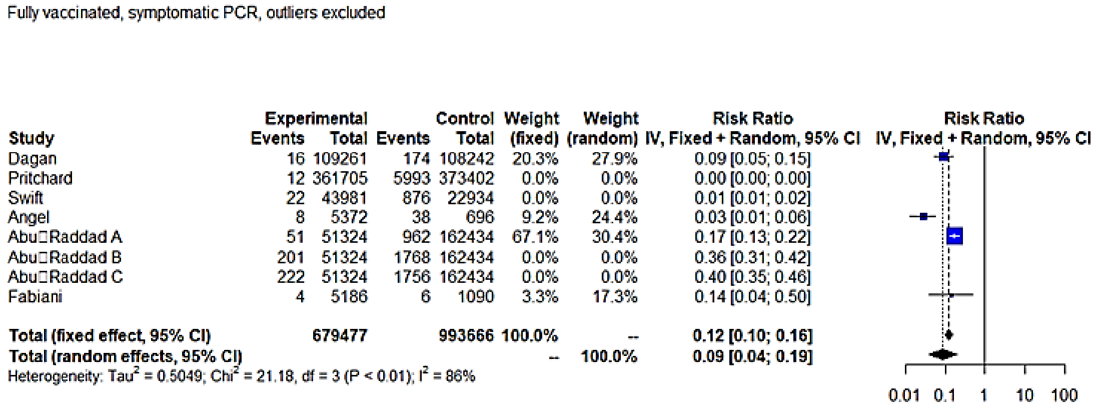


**Fig.23a) Influence analysis.** Symptomatic positive Sars-Cov-2 PCR, fully vaccinated. “Pritchard” (red dot) resulted as the most influential study as it exceeded the cut-offs defined by Viechtbauerand Cheung in any subplot.[21]

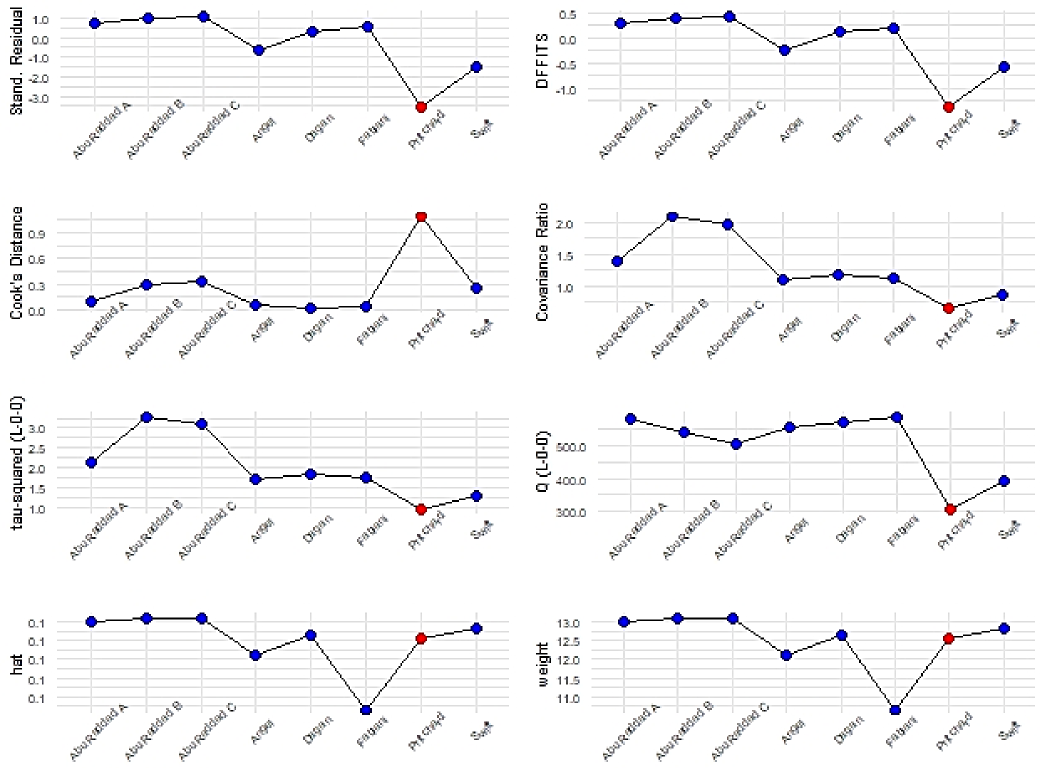

**Fig.23b) Baujat plot**. Fully vaccinated, symptomatic PCR. Contribute to overall heterogeneity, Q (x) and effect size (y). “Pritchard” and “Swift” studies contributed to the pooled imbalance. “Abu Raddad C” was the most influential on the pooled effect size.

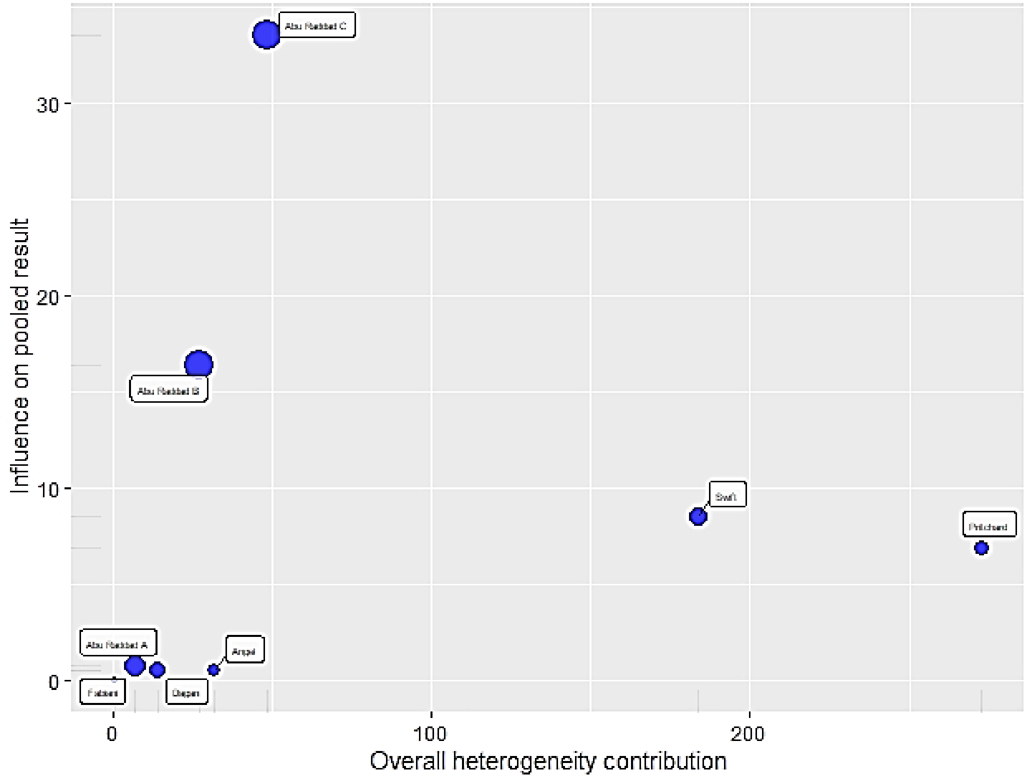


**Fig.24) Leave one out analysis.** Fully vaccinated, symptomatic PCR. The first plot displayed the change on the I2 heterogeneity attained by removing one study at each step. “Pritchard” was the most influential on the pooled imbalance, while “Abu Raddad” affected the overall RR estimate.

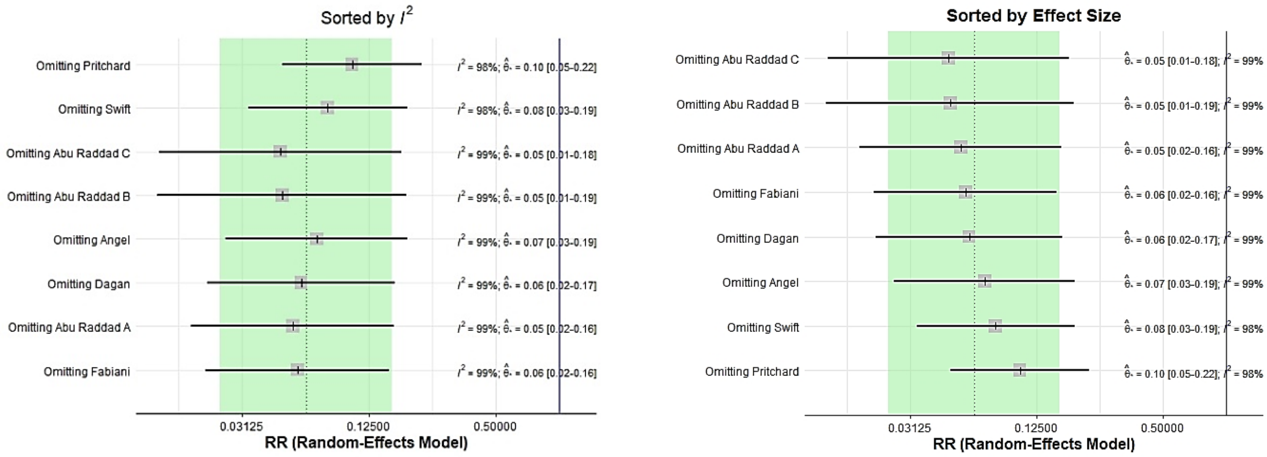

**Fig.25) GOSH Diagnostics.** Fully vaccinated, symptomatic PCR. GOSH plots did not show any pattern, The effect size distribution curve was symmetric. The heterogeneity curve (I2) was right skewed with a sharp peak. The three algorithms, k -means, DBSCAN and the Gaussian Mixture Model, detected up to eight clusters which might potentially contribute to the overall heterogeneity, including “Pritchard”(5).

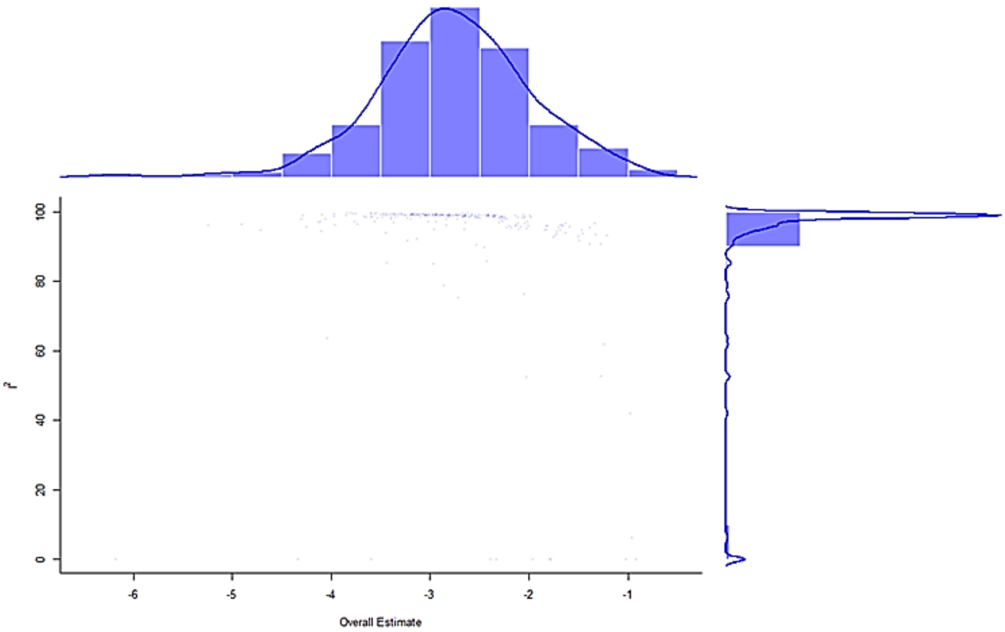


| GOSH Diagnostics |
| --- |
| - Number of K-means clusters detected: 3 |
| - Number of DBSCAN clusters detected: 4 |
| - Number of GMM clusters detected: 8 |
| **Identification of potential outliers** |
| - K-means: Study 8, Study 2 |
| - DBSCAN: Study 2 |
| - Gaussian Mixture Model: Study 2 |

### Annex 4

### A4.1 Subgroup analysis

**Tab.10)** Random and Mixed model, any positive PCR RR

| Model | Subgroups | | N | Results for subgroups | | | Between-groups heterogeneity | | |
|---|---|---|---|---|---|---|---|---|---|
| | | | | RR [95%-CI] | I2 | τ2 | Q | df(Q) | p-value |
| **Partially vaccinated, any positive PCR** | | | | | | | | | |
| Mixed model | Vaccine | BNT162b2 | 16 | 0.2781 [0.1800; 0.4298] | 99.60% | 0.7443 | 6.07 | 3 | 0.1083 |
| | | ChAdOx1 | 2 | 0.2889 [0.0871; 0.9579] | 98.90% | 0.7443 | | | |
| | | BNT162b2/mRNA-1273 | 3 | 0.3933 [0.1463; 1.0575] | 98.80% | 0.7443 | | | |
| | | BNT162b2/ChAdOx1 | 1 | 0.0357 [0.0066; 0.1939] | -- | -- | | | |
| | Quality | NOS ≤6 | 15 | 0.2985 [0.1986; 0.4485] | 99.60% | 0.6113 | 0.21 | 1 | 0.6438 |
| | | NOS >6 | 7 | 0.2131 [0.0543; 0.8367] | 99.70% | 3.3748 | | | |
| | Age | < 69 years | 15 | 0.2394 [0.1259; 0.4554] | 99.80% | 1.5669 | 0.74 | 1 | 0.3909 |
| | | ≥ 69 years | 7 | 0.3317 [0.2278; 0.4828] | 98.60% | 0.2424 | | | |
| | Lineage | B.1.1.7 | 8 | 0.1895 [0.0917; 0.3918] | 99.70% | 1.039 | 3.21 | 3 | 0.3606 |
| | | B.1.1.7/non B.1.1.7 | 5 | 0.2147 [0.0875; 0.5266] | 99.70% | 1.039 | | | |
| | | not specified | 8 | 0.3615 [0.1763; 0.7415] | 97.10% | 1.039 | | | |
| | | B.1.351 | 1 | 0.9080 [0.1231; 6.6974] | -- | -- | | | |
| Random Effect model | Vaccine | BNT162b2 | 16 | 0.2781[0.1794;0.4310] | -- | 0.7549 | 116.47 | 3 | **< 0.0001** |
| | | ChAdOx1 | 2 | 0.2885[0.1256;0.6629] | -- | 0.3565 | | | |
| | | BNT162b2/mRNA-1273 | 3 | 0.3942[0.1210;1.2838] | -- | 1.0692 | | | |
| | | BNT162b2/ChAdOx1 | 1 | 0.0357[0.0323;0.0393] | -- | -- | | | |
| | Quality | NOS ≤6 | 15 | 0.2985[0.1986;0.4485] | -- | 0.6113 | 0.21 | 1 | 0.6438 |
| | | NOS >6 | 7 | 0.2131[0.0543;0.8367] | -- | 3.3748 | | | |
| | Age | < 69 years | 15 | 0.2394[0.1259;0.4554] | -- | 1.5669 | 0.74 | 1 | 0.3909 |
| | | ≥ 69 years | 7 | 0.3317[0.2278;0.4828] | -- | 0.2424 | | | |
| | Lineage | B.1.1.7 | 8 | 0.1894[0.0946;0.3792] | -- | 0.9447 | 36.34 | 3 | **< 0.0001** |
| | | B.1.1.7/non B.1.1.7 | 5 | 0.2148[0.0750;0.6147] | -- | 1.4303 | | | |
| | | not specified | 8 | 0.3618[0.2039;0.6422] | -- | 0.6506 | | | |
| | | B.1.351 | 1 | 0.9080[0.8717;0.9458] | -- | -- | | | |
| **Fully vaccinated, any positive PCR** | | | | | | | | | |
| Mixed model | Vaccine | BNT162b2 | 11 | 0.0628 [0.0261; 0.1511] | 98.40% | 2.1113 | 3.21 | 3 | 0.3609 |
| | | BNT162b2/mRNA-1273 | 4 | 0.0639 [0.0144; 0.2844] | 98.90% | 2.1113 | | | |
| | | BNT162b2/ChAdOx1 | 1 | 0.0062 [0.0004; 0.1072] | -- | -- | | | |
| | | Ad26.COV2.S | 1 | 0.2338 [0.0109; 5.0306] | -- | -- | | | |
| | Quality | NOS ≤6 | 11 | 0.0785 [0.0383; 0.1609] | 98.50% | 1.3935 | 1.66 | 1 | 0.1973 |
| | | NOS >6 | 6 | 0.0332 [0.0111; 0.0991] | 95.90% | 1.2835 | | | |
| | Age | < 69 years | 13 | 0.0479 [0.0182; 0.1256] | 99.30% | 3.0726 | 2.29 | 1 | 0.1304 |
| | | ≥ 69 years | 4 | 0.1200 [0.0597; 0.2412] | 65.00% | 0.2931 | | | |
| | Lineage | B.1.1.7 | 4 | 0.0786 [0.0190; 0.3240] | 95.90% | 2.0252 | 3.27 | 3 | 0.3515 |
| | | B.1.1.7/non B.1.1.7 | 5 | 0.0285 [0.0081; 0.1005] | 98.60% | 2.0252 | | | |
| | | not specified | 7 | 0.0632 [0.0206; 0.1934] | 97.70% | 2.0252 | | | |
| | | B.1.351 | 1 | 0.4027 [0.0247; 6.5721] | -- | -- | | | |
| Random Effect model | Vaccine | BNT162b2 | 11 | 0.0631 [0.0281; 0.1416] | -- | 1.7775 | 67.41 | 3 | **< 0.0001** |
| | | BNT162b2/mRNA-1273 | 4 | 0.0646 [0.0092; 0.4548] | -- | 3.7392 | | | |
| | | BNT162b2/ChAdOx1 | 1 | 0.0062 [0.0049; 0.0078] | -- | -- | | | |
| | | Ad26.COV2.S | 1 | 0.2338 [0.0745; 0.7337] | -- | -- | | | |
| | Quality | NOS ≤6 | 11 | 0.0785 [0.0383; 0.1609] | -- | 1.3935 | 1.66 | 1 | 0.1973 |
| | | NOS >6 | 6 | 0.0332 [0.0111; 0.0991] | -- | 1.6474 | | | |
| | Age | < 69 years | 13 | 0.0479 [0.0182; 0.1256] | -- | 3.0726 | 2.29 | 1 | 0.1304 |
| | | ≥ 69 years | 4 | 0.1200 [0.0597; 0.2412] | -- | 0.2931 | | | |
| | Lineage | B.1.1.7 | 4 | 0.0810 [0.0323; 0.2031] | -- | 0.817 | 33.99 | 3 | **< 0.0001** |
| | | B.1.1.7/non B.1.1.7 | 5 | 0.0284 [0.0081; 0.0996] | -- | 1.9969 | | | |
| | | not specified | 7 | 0.0634 [0.0151; 0.2658] | -- | 3.4732 | | | |
| | | B.1.351 | 1 | 0.4027 [0.3533; 0.4592] | -- | -- | | | |
| **At least one dose, any positive PCR** | | | | | | | | | |
| Mixed model | Vaccine | BNT162b2 | 12 | 0.1473 [0.0946; 0.2292] | 99.60% | 0.5994 | 2.24 | 2 | 0.3256 |
| | | BNT162b2/mRNA-1273 | 4 | 0.2622 [0.1215; 0.5656] | 98.50% | 0.5994 | | | |
| | | ChAdOx1 | 2 | 0.1061 [0.0350; 0.3214] | 83.10% | 0.5994 | | | |
| | Quality | NOS ≤6 | 16 | 0.1793 [0.1206; 0.2666] | 99.40% | 0.6388 | 2.35 | 1 | 0.1252 |

| Model | | Subgroups | N | RR [95%-CI] | I2 | τ2 | Q | df(Q) | p-value |
|---|---|---|---|---|---|---|---|---|---|
| | | NOS >6 | 2 | 0.0711 [0.0233; 0.2165] | 99.90% | 0.6388 | | | |
| | Age | < 69 years | 12 | 0.1325 [0.0879; 0.1998] | 99.30% | 0.5076 | 2.53 | 1 | 0.1116 |
| | | ≥ 69 years | 6 | 0.2395 [0.1311; 0.4375] | 99.80% | 0.5585 | | | |
| | Lineage | B.1.1.7 | 4 | 0.1523 [0.0690; 0.3362] | 99.80% | 0.6484 | 2.2 | 2 | 0.3325 |
| | | B.1.1.7/non B.1.1.7 | 5 | 0.1059 [0.0514; 0.2184] | 95.00% | 0.6484 | | | |
| | | not specified | 9 | 0.2081 [0.1224; 0.3538] | 99.80% | 0.6484 | | | |
| Random Effect model | Vaccine | BNT162b2 | 12 | 0.1473 [0.0939; 0.2310] | -- | 0.6218 | 3.68 | | 0.1588 |
| | | BNT162b2/mRNA-1273 | 4 | 0.2646 [0.1402; 0.4994] | -- | 0.4039 | | | |
| | | ChAdOx1 | 2 | 0.1099 [0.0551; 0.2190] | -- | 0.2114 | | | |
| | Quality | NOS ≤6 | 16 | 0.1800 [0.1296; 0.2499] | -- | 0.432 | 0.2 | | 0.6541 |
| | | NOS >6 | 2 | 0.0696 [0.0011; 4.3786] | -- | 8.9219 | | | |
| | Age | < 69 years | 12 | 0.1325 [0.0879; 0.1998] | -- | 0.5076 | 2.53 | | 0.1116 |
| | | ≥ 69 years | 6 | 0.2395 [0.1311; 0.4375] | -- | 0.5585 | | | |
| | Lineage | B.1.1.7 | 4 | 0.1518 [0.0602; 0.3827] | -- | 0.8863 | 3.13 | | 0.2096 |
| | | B.1.1.7/non B.1.1.7 | 5 | 0.1070 [0.0629; 0.1822] | -- | 0.3366 | | | |
| | | not specified | 9 | 0.2081 [0.1250; 0.3464] | -- | 0.5975 | | | |

#### **Tab.11)** Random and Mixed model, symptomatic PCR RR

| **Partially vaccinated, symptomatic PCR** | | | | | | | | | |
|---|---|---|---|---|---|---|---|---|---|
| **Model** | **Subgroups** | | **N** | **Results for subgroups** | | | **Between-groups heterogeneity** | | |
| | | | | **RR [95%-CI]** | **I2** | **τ2** | **Q** | **df(Q)** | **p-value** |
| Mixed model | Vaccine | BNT162b2 | 6 | 0.2488 [0.1035; 0.5980] | 96.20% | 1.1044 | 4.96 | 3 | 0.1748 |
| | | ChAdOx1 | 1 | 0.3441 [0.0395; 3.0002] | -- | -- | | | |
| | | BNT162b2/ChAdOx1 | 1 | 0.0270 [0.0034; 0.2134] | -- | -- | | | |
| | | BNT162b2/mRNA-1273 | 1 | 0.5552 [0.0697; 4.4200] | -- | -- | | | |
| | Quality | NOS ≤6 | 6 | 0.3007 [0.0875; 1.0332] | 93.60% | 2.2707 | 0.76 | 1 | 0.3823 |
| | | NOS >6 | 3 | 0.1172 [0.0211; 0.6514] | 99.50% | 2.2707 | | | |
| | Age | < 69 years | 7 | 0.1860 [0.0468; 0.7385] | 99.60% | 3.384 | 1.08 | 1 | 0.2993 |
| | | ≥ 69 years | 2 | 0.3962 [0.2736; 0.5737] | 0.00% | 0 | | | |
| | Lineage | B.1.1.7 | 3 | 0.2498 [0.0226; 2.7633] | 89.10% | 4.4369 | 0.13 | 2 | 0.9384 |
| | | B.1.1.7/non B.1.1.7 | 2 | 0.1367 [0.0074; 2.5364] | 99.90% | 4.4369 | | | |
| | | not specified | 4 | 0.2504 [0.0308; 2.0320] | 94.10% | 4.4369 | | | |
| Random Effect model | Vaccine | BNT162b2 | 6 | 0.2488 [0.1035; 0.5980] | 96.20% | 1.1044 | 437.47 | 3 | **< 0.0001** |
| | | ChAdOx1 | 1 | 0.3441 [0.1763; 0.6715] | -- | -- | | | |
| | | BNT162b2/mRNA-1273 | 1 | 0.0270 [0.0231; 0.0317] | -- | -- | | | |
| | | BNT162b2/ChAdOx1 | 1 | 0.5552 [0.4336; 0.7111] | -- | -- | | | |
| | Quality | NOS ≤6 | 6 | 0.3062 [0.1465; 0.6400] | 93.60% | 0.7447 | 0.69 | 1 | 0.4071 |
| | | NOS >6 | 3 | 0.1173 [0.0137; 1.0027] | 99.50% | 3.5694 | | | |
| | Age | < 69 years | 7 | 0.1860 [0.0468; 0.7385] | 99.60% | 3.384 | 1.08 | 1 | 0.2993 |
| | | ≥ 69 years | 2 | 0.3962 [0.2736; 0.5737] | 0.00% | 0 | | | |
| | Lineage | B.1.1.7 | 3 | 0.2482 [0.1001; 0.6152] | 89.10% | 0.5695 | 0.13 | 2 | 0.936 |
| | | B.1.1.7/non B.1.1.7 | 2 | 0.1367 [0.0057; 3.2685] | 99.90% | 5.242 | | | |
| | | not specified | 4 | 0.2526 [0.0776; 0.8228] | 94.10% | 1.3278 | | | |
| **Fully vaccinated, symptomatic PCR** | | | | | | | | | |
| Mixed model | Vaccine | BNT162b2 | 6 | 0.1603 [0.0966; 0.2661] | 94.90% | 0.3245 | 48.24 | 2 | **< 0.0001** |
| | | BNT162b2/mRNA-1273 | 1 | 0.0131 [0.0040; 0.0432] | -- | -- | | | |
| | | BNT162b2/ChAdOx1 | 1 | 0.0021 [0.0006; 0.0072] | -- | -- | | | |
| | Quality | NOS ≤6 | 5 | 0.2156 [0.1279; 0.3633] | 93.00% | 0.2922 | 52.21 | 1 | **< 0.0001** |
| | | NOS >6 | 3 | 0.0087 [0.0043; 0.0174] | 94.60% | 0.2922 | | | |
| | Lineage | B.1.1.7 | 2 | 0.0684 [0.0025; 1.8450] | 94.80% | 5.5699 | 0.69 | 3 | 0.8757 |
| | | B.1.1.7/non B.1.1.7 | 3 | 0.0427 [0.0029; 0.6248] | 99.40% | 5.5699 | | | |
| | | not specified | 2 | 0.0413 [0.0015; 1.1598] | 91.80% | 5.5699 | | | |
| | | B.1.351 | 1 | 0.3598 [0.0035; 36.8088] | -- | -- | | | |
| Random Effect model | Vaccine | BNT162b2 | 6 | 0.1603 [0.0966; 0.2661] | 94.90% | 0.3245 | 129.86 | 3 | **< 0.0001** |
| | | BNT162b2/mRNA-1273 | 1 | 0.0131 [0.0086; 0.0200] | -- | -- | | | |
| | | BNT162b2/ChAdOx1 | 1 | 0.0021 [0.0012; 0.0036] | -- | -- | | | |
| | Quality | NOS ≤6 | 5 | 0.2214 [0.1435; 0.3416] | 93.00% | 0.1911 | 18.17 | 1 | **< 0.0001** |
| | | NOS >6 | 3 | 0.0089 [0.0022; 0.0366] | 94.60% | 1.4624 | | | |
| | Lineage | B.1.1.7 | 2 | 0.0701 [0.0118; 0.4152] | 94.80% | 1.565 | 8.61 | 3 | **0.035** |
| | | B.1.1.7/non B.1.1.7 | 3 | 0.0426 [0.0022; 0.8254] | 99.40% | 6.8062 | | | |
| | | not specified | 2 | 0.0396 [0.0039; 0.4023] | 91.80% | 2.5779 | | | |
| | | B.1.351 | 1 | 0.3598 [0.3111; 0.4162] | | -- | | | |

#### A4.2 Publication bias

**Fig. 26) Funnel plot and Eggers' test.** Any positive PCR (a, b, c). Symptomatic PCR (d, e). The contour-enhanced funnels included three colours signifying the significance level into which the effects size of each study fell. The dots' distribution was overall quite sparse and concentrated at the upper half of the funnel except "Tenforde" at the bottom.

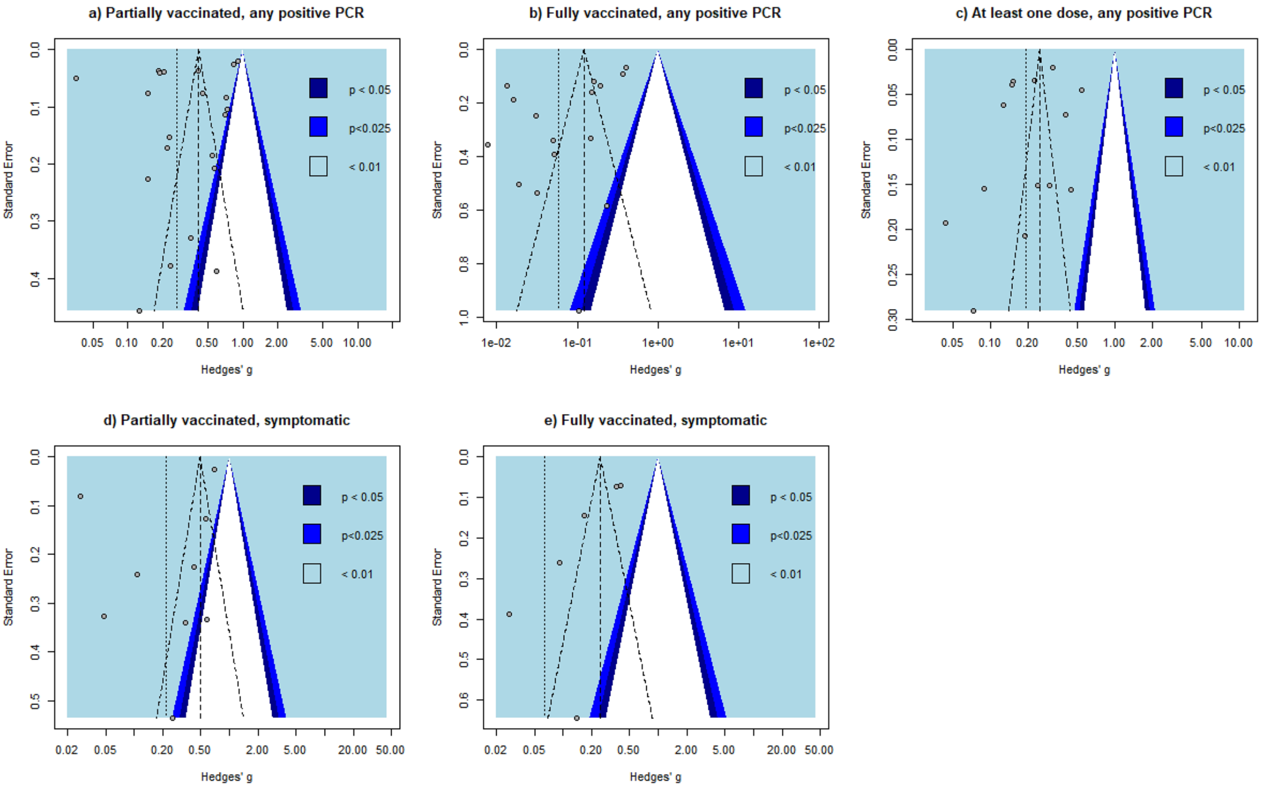


| Eggers' test of the intercept | | | |
|---|---|---|---|
| **intercept** | **95% CI** | **t** | **p** |
| **a) Any positive PCR partially vaccinated** | | | |
| -8.868 | -20.25 - 2.51 | -1.527 | 0.1423509 |
| **b) Any positive PCR fully vaccinated** | | | |
| -6.792 | -14.78 - 1.2 | -1.667 | 0.1163403 |
| **c) Any positive PCR at least one dose** | | | |
| -3.843 | -11.65 - 3.96 | -0.965 | 0.3537481 |
| **d) Symptomatic positive PCR partially vaccinated** | | | |
| -6.858 | -18.21 - 4.5 | -1.184 | 0.2751005 |
| **e) Symptomatic positive PCR fully vaccinated** | | | |
| -11.271 | -18.76 - -3.78 | -2.948 | **0.02567738** |
